# Estimating tau onset age from tau PET imaging in two longitudinal cohorts using sampled iterative local approximation

**DOI:** 10.64898/2026.04.01.26349872

**Authors:** Tobey J Betthauser, Jordan P Teague, Hailey Bruzzone, Margo B Heston, William Coath, Jacob D Morse, Elena L Ruiz de Chavez, Finnuella J Carey, Ruvini Navaratna, Karly A Cody, Rebecca E Langhough

**Affiliations:** Wisconsin Alzheimer’s Disease Research Center, University of Wisconsin School of Medicine and Public Health, Madison, WI, USA; Department of Medicine, University of Wisconsin-Madison School of Medicine and Public Health, Madison, WI, USA; Department of Medical Physics, University of Wisconsin-Madison School of Medicine and Public Health, Madison, WI, USA; Center for Health Disparities Research, University of Wisconsin School of Medicine and Public Health, Madison, WI, USA; Wisconsin Alzheimer’s Institute, University of Wisconsin School of Medicine and Public Health, Madison, WI, USA; Department of Neurology and Neurological Sciences, Stanford University, Stanford, CA, USA

**Keywords:** Tau, PET, temporal modeling, multicohort, Alzheimer’s, SILA

## Abstract

Understanding the time course of Alzheimer’s disease biomarkers of amyloid and tau pathology and their temporal relation to clinical symptoms is key to identifying optimal windows for disease intervention and planning future drug trials. The goal of this work was to determine the extent to which Sampled Iterative Local Approximation (SILA), an algorithm extensively validated for amyloid PET, is capable of modeling longitudinal tau (T) PET trajectories and estimating person-level tau positivity onset ages in two commonly analyzed brain regions and two tracers from two different cohorts

**Methods:** 385 participants from the Alzheimer’s Disease Neuroimaging Initiative (ADNI; mean (SD) age = 73.4 (7.3) years) with longitudinal flortaucipir tau PET and 288 participants from the Wisconsin Registry for Alzheimer’s Prevention and Wisconsin Alzheimer’s Disease Research Center (collectively referred to as WISC; mean (SD) age = 67.4 (6.7) years) with longitudinal MK-6240 tau PET were included in the study. Standard uptake value ratios (SUVRs) in the entorhinal cortex and a meta-temporal ROI were modeled with SILA separately, for each cohort and region. Forward and backward SUVR and T+/- prediction were characterized with ten-fold cross-validation and in-sample validation techniques. Accuracy of estimated T+ onset ages (ETOA) was characterized in T- to T+ converters. Differences in ETOA were tested between *APOE-e4* carriers and non-carriers, as well as differences in time T+ between levels of cognitive impairment.

**Results:** SILA was able to accurately estimate retrospective change in tau SUVR in the meta-temporal region regardless of age, sex, *APOE-e4* carriage, tau SUVR, and dementia (p >0.05) whereas dementia was associated with model residuals in entorhinal cortex (p ≤0.05; ADNI). In subsets of observed T- to T+ converters, the difference between “observed” and estimated meta-temporal T+ onset age [95% CI] was 0.12 [-0.27, 0.52] years for ADNI and -0.09 [0.93, 0.74] years for WISC. ETOA was significantly earlier, and odds of SILA-estimated T+ status were higher amongst *APOE-e4* carriers (p <0.05) and those with dementia (p <0.05).

**Conclusions:** Our results suggest SILA can be used to accurately model longitudinal tau PET trajectories and retrospectively estimate individual T+ onset ages in the meta-temporal region. The accuracy of SILA time estimates in entorhinal cortex worsened amongst those with dementia in ADNI suggesting entorhinal cortex may only be suitable for studying the temporal progression of tau during the preclinical time frame.

## 1. Introduction

Alzheimer’s disease (AD) biomarkers have played transformative roles in characterizing disease pathophysiology and advancing clinical trials including the recent discovery of new disease modifying therapies (Jack et al., 2018; Johnson et al., 2013; Rabinovici et al., 2019; Rabinovici & La Joie, 2023). Overwhelmingly, observational studies suggest that AD biomarkers become abnormal 20 years or more before clinical symptoms (Betthauser et al., 2022; Jagust et al., 2021; Schindler et al., 2021; Villemagne et al., 2013) and tend to follow a stereotypic sequence beginning with amyloid positivity (A+), tau positivity (T+), and subsequent neurodegenerative changes and clinical impairment (Bateman et al., 2011; Jack et al., 2013). This 20+ year disease process poses a challenge for characterizing the full course of disease since it is uncommon to have longitudinal observation of individuals spanning the transition from the earliest detectable biomarker changes into clinical impairment. This necessitates the development of validated methods that utilize shorter longitudinal observations of individuals (e.g., 5-10 years of longitudinal follow-up) to piece together the full disease course.

Several recent studies have used and/or tested methods for modeling the progression of AD biomarkers using multiple biomarkers simultaneously (e.g., progression scores) (Cogswell et al., 2024; Jedynak et al., 2012), or using individual biomarkers to define biomarker-specific timelines (Betthauser et al., 2022; Bilgel et al., 2016; Koscik et al., 2020; Schindler et al., 2021; Therneau et al., 2021; Whittington et al., 2018). Many of the latter studies have focused on modeling longitudinal amyloid positron emission tomography (PET) trajectories to define the preclinical disease timeline (i.e., the time from A+ to clinical impairment). These studies consistently demonstrate that amyloid PET accumulation follows a highly predictable trajectory in sporadic AD once detectable biomarker levels are reached, regardless of common AD risk/protective factors like sex, age, and apolipoprotein-E (*APOE*) genotype. This consistency in amyloid accumulation can be exploited to generate population/sample-level amyloid accumulation trajectory curves trained on longitudinal cohort data, which can then be used to reliably estimate when individuals became A+ (i.e., “estimated A+ onset age”; EAOA), even when the transition from A- to A+ is not directly observed (Betthauser et al., 2022; Bilgel et al., 2016; Koscik et al., 2020; Schindler et al., 2021). A+ onset ages and A+ onset-aligned amyloid curves have proven useful in several ways, including: 1) establishing an easily interpretable disease time metric (calculated as measurement age minus EAOA; referred to as amyloid chronicity, duration, time, or clock); 2) enabling accurate estimation of amyloid burden for study visits that occurred prior to the availability of a PET scan; 3) illustrating the heterogeneity in A+ onset ages; 4) facilitating investigation of factors related to A+ risk and A+ onset age; and 5) deeper characterization of factors influencing time between A+ onset and dementia.

While methods have been extensively validated for modeling the amyloid accumulation, little work has been done to validate methods for modeling longitudinal tau PET trajectories and subsequent estimation of individual T+ onset ages. In contrast to amyloid, PET-measured tau positivity is expected to be more temporally proximal to the onset of clinical symptoms with a spatiotemporal progression typically beginning in the entorhinal cortex and later spreading to adjacent temporal and further neocortical regions (Braak & Braak, 1991; Cody et al., 2024; Pascoal et al., 2021). Higher tau burden, particularly outside of the medial temporal lobe, has been shown to have a stronger association with cognitive decline and worsening of clinical symptoms compared to amyloid (Jack et al., 2024; Therriault et al., 2022). Validated tau PET modeling methods are needed to help facilitate the study of timing between biomarker events (e.g., A+ to T+, p-Tau217+ to T+), timing from T+ to dementia onset, and factors that may slow or hasten these progressions. Additionally, methods that could reliably estimate tau PET standard uptake value ratios (SUVR) prior to first tau PET scan would help bridge the time gap in many longitudinal neuroimaging studies between the first plasma observations, amyloid PET scans, and initial tau PET scans (often occurring 5-10 years later due to the later discovery of tau PET).

In this work, we validate sampled iterative local approximation (SILA), a temporal modeling method that has been extensively validated for amyloid PET, to estimate longitudinal tau PET trajectories and individualized T+ onset ages. We present separate analyses of two cohorts using different tau PET tracers to investigate the following: **Aim 1** focuses on evaluating the validity of SILA-modeled tau PET trajectories using the following criteria: 1a) consistency of SILA-modeled tau trajectories across folds using 10-fold cross-validation 1b) accuracy of forward and backward SUVR predictions based on single reference scans, and 1c) precision in predicting T+ onset age compared to observed T+ onset age. **Aim 2** explores how SILA-derived T+ rates, T+ time, and T+ onset age relate to common AD risk factors by examining: 2a) tau positivity rates in relation to *APOE-ε4* status and dementia diagnosis, and 2b) estimated tau onset age (ETOA) by *APOE-ε4* status and T+ time by dementia status.

## 2. Methods

### 2.1 Study Cohorts and Inclusion Criteria

Analysis was performed using tau PET data collected by the Alzheimer’s Disease Neuroimaging Initiative (ADNI) as well as data from the combined Wisconsin Alzheimer’s Disease Research Center (WADRC) and the Wisconsin Registry for Alzheimer’s Prevention (WRAP); collectively referred to as WISC. Participants with two or more tau PET scans available at the time of analysis were included. Detailed descriptions of these cohorts are provided in the following references (Johnson et al., 2018; Mueller et al., 2005). Briefly, ADNI was launched in 2003 as a public-private partnership, led by Principal Investigator Michael W. Weiner, MD. The primary goal of ADNI has been to test whether serial magnetic resonance imaging (MRI), PET, other biological markers, and clinical and neuropsychological assessment can be combined to measure the progression of mild cognitive impairment (MCI) and early AD. For up-to-date information, see www.adni-info.org. WRAP started in 2001, led initially by Principal Investigator, Mark Sager, MD, and since 2014, by Principal Investigator, Sterling Johnson, PhD. The Wisconsin ADRC was established in 2009 led by Principal Investigator, Sanjay Asthana, MD. Both WISC cohorts are designed to characterize how lifestyle, genetic, and other risks associate with preclinical changes and AD progression. Both are enriched for parental history of AD. Source studies were conducted under their respective regulatory agencies including written informed consent. Secondary analyses for this work were approved and conducted under the University of Wisconsin Institutional Review Board in accordance with the Declaration of Helsinki.

### 2.2 Clinical Characterization

The Clinical Dementia Rating (CDR) (Morris, 1993) was used to establish clinical status as unimpaired (CDR global = 0), very mild dementia (CDR global = 0.5), and mild, moderate or severe dementia (CDR global = 1, 2, 3, respectively). We used CDR instead of clinical diagnosis because the CDR was administered the same across cohorts and is frequently used as a primary or secondary outcome for clinical trials (Sims et al., 2023; van Dyck et al., 2023). CDR closest in time to the last tau PET scan was used in analyses.

### 2.3 ADNI Tau PET image acquisition and quantification

Data for [^18^F]flortaucipir (FTP) analyses were processed by UC-Berkeley and downloaded from Laboratory of NeuroImaging Image & Data Archive. Details regarding FTP PET image acquisition and processing are provided in the following references (Baker, Lockhart, et al., 2017; Baker, Maass, et al., 2017). FTP data were acquired 80-100 minutes after bolus injection in four, 5-minute frames. Prior to image processing, data were converted to NIfTI format, inter-frame aligned and smoothed to a uniform resolution using scanner-specific smoothing kernels derived from Hoffman phantom studies. Individual PET frames were summed and rigidly registered to T1-weighted MRI that have undergone FreeSurfer v7.1.1 anatomical parcellation. Regional SUVRs were extracted from FreeSurfer regions of interest (ROIs) using the inferior cerebellum gray matter as a reference region.

### 2.4 WISC Tau PET image acquisition and quantification

[^18^F]MK-6240 tau PET images for the WISC cohorts were acquired and quantified as previously described (Betthauser et al., 2018). Briefly, [^18^F]MK-6240 images were acquired on either a Siemens Biograph Horizon PET/CT or Siemens ECAT EXACT HR+ tomograph after 185 or 370 MBq bolus injection. Scan acquisition includes data corresponding to 70-90 minutes post-injection. PET and T1-weighted MRI were processed using SPM12 and MATLAB. T1-weighted MRI were tissue class segmented and spatially normalized to MNI-152 space. ROIs from the Harvard-Oxford atlas were inverse warped to subject T1-weighted image space and limited to voxels with gray matter (GM) probabilities >0.3. PET time series were smoothed, interframe aligned, and co-registered to T1-weighted MRI. SUVR (inferior cerebellum reference region) was quantified in the entorhinal cortex (EC, corresponding to the anterior parahippocampal gyrus Harvard-Oxford ROI) and in a meta-temporal (MT) ROI (amygdala, parahippocampal gyrus, temporal fusiform cortex, inferior temporal gyrus, and middle temporal gyrus) (Jack et al., 2017) using a volume-weighted SUVR average across left and right brain regions.

### 2.5 Regional T+ thresholds

To ensure methodological consistency for regional tau positivity (T+/-) across tracers/ cohorts, Gaussian mixture models (GMM) were used to determine region- and cohort-specific tau positivity thresholds (fitgmdist, MATLAB). For each combination of tracer and ROI, a GMM was applied to baseline SUVR, assuming two classes represented in the data. We defined T+ thresholds as the mean plus two standard deviations (SD) from the lower GMM distribution. This resulted in the following T+ thresholds: ADNI EC SUVR >1.32; ADNI MT SUVR >1.37; WISC EC SUVR >1.27; WISC MT SUVR >1.32.

### 2.6 SILA implementation

#### 2.6.1 - Model training

The SILA method is described in detail in the following reference (Betthauser et al., 2022). Briefly, SILA uses a discrete sampling approach to generate a numeric curve representing the mean rate of change at evenly spaced, discrete SUVR values spanning the minimum and maximum values in the input data. This numeric curve is then smoothed and integrated using Euler’s method to generate a numeric SUVR vs. time curve. To anchor the curve to a meaningful event, the time axis is defined such that 0 years corresponds to the user-specified T+ threshold. All SILA models used a step size of 0.25 years and max of 100 iterations.

#### 2.6.2 SILA Individual T+ time, T+ age, and SUVR estimation

After model training, T+ time and T+ age (i.e., ETOA) are estimated for each participant based on one or all reference observations (i.e., tau PET scan(s)). For the reference observation, the numeric SUVR vs. T+ time curve defined in 2.6.1 is solved for time given an input observed SUVR value. This estimated T+ time is subtracted from age at reference observation to estimate T+ age. To generate estimated SUVRs at different time points, the numeric SUVR vs. T+ time curve from 2.6.1 is solved for SUVR at the estimated T+ time plus or minus time between the reference observation and another timepoint (e.g., a previous or later tau PET scan). Positive change in time corresponds to an observation after the reference observation (i.e., future/prospective) whereas negative change in time corresponds to an observation before the reference observation (i.e., past/retrospective).

### 2.7 Statistical Analysis

A flow diagram for model training and analysis is provided in Figure 1. SILA model training, estimation, and residual calculations between observed and estimated SUVR were performed using MATLAB v2021a software (9.10.0.1649659) on a Dell Precision 7920 desktop with an Intel Xeon Gold 6248R processor (3.0 GHz base clock, 4.0 GHz Turbo, 24 Cores), 128 GB of random-access memory (DDR4, 2933 MHz) and Windows 10 Enterprise (Version 22H2). Statistical analyses were performed in R v4.4.2. Aim 1 used 10-fold cross-validation and whole-sample training to assess SILA’s prediction of tau PET, with sub-analyses on model convergence (1a), SUVR and T+/- prediction accuracy (1b), and estimated T+ onset age (1c). Aim 2 examined T+ rates (2a) and the relationship of T+ time and tau onset with AD risk factors (2b)

**Figure 1:**
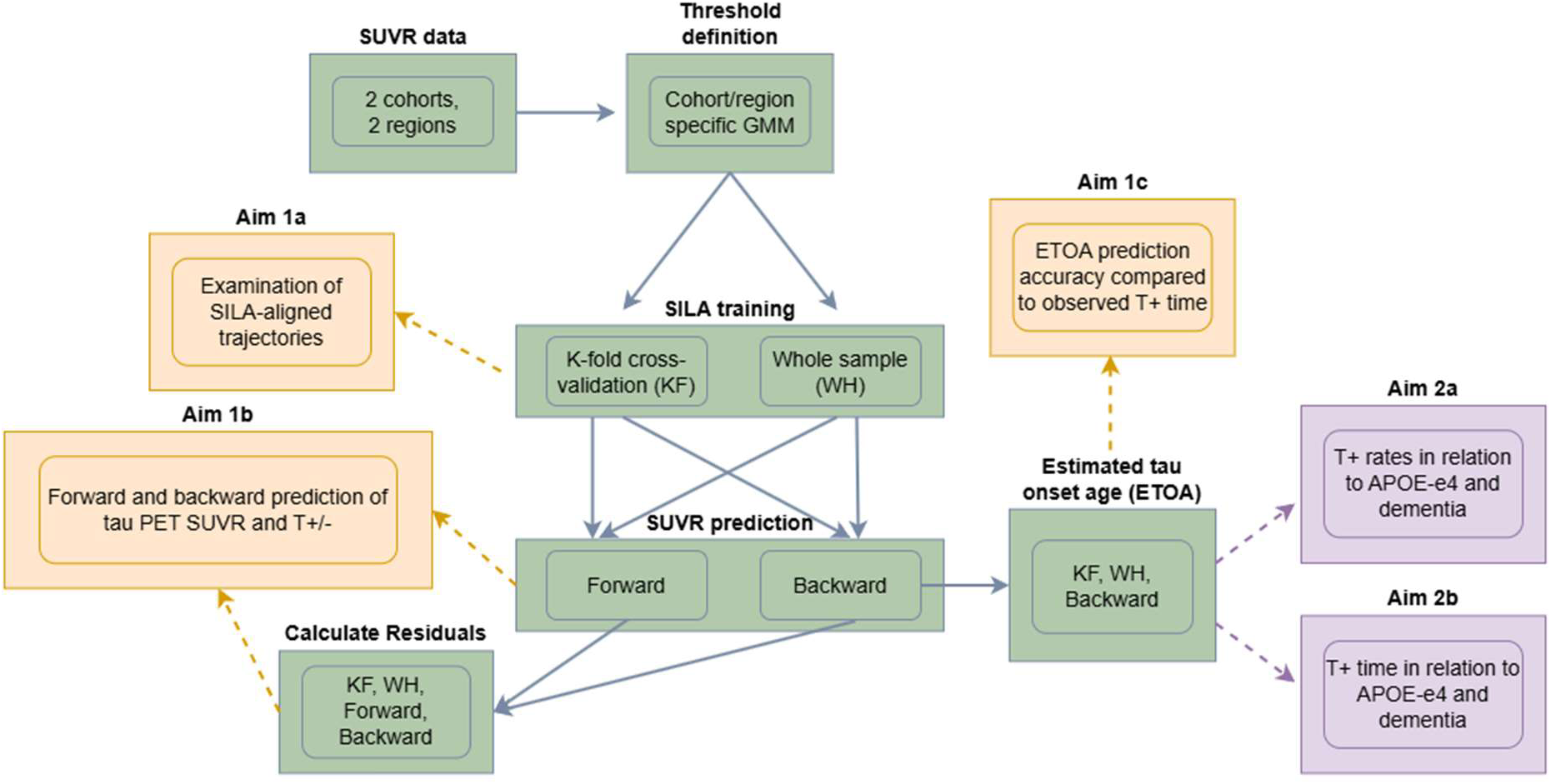
Analysis flow diagram. Flow diagram depicts how cohort and ROI SUVR data were used to validate the SILA algorithm. *APOE-*e4 = apolipoprotein e4, ETOA = estimated tau onset age, GMM = Gaussian mixture modeling, KF = k-fold cross-validation, PET = positron emission tomography, SILA = sampled iterative local approximation, SUVR = standard uptake value ratio, WH = whole sample

#### 2.7.1 Partitioning Tau PET data for 10-fold cross validation

For Aim 1, data for each cohort were partitioned into 10 folds using the MATLAB *cvpartition* function. To ensure each fold contained individuals accumulating tau for model training (i.e., those who were T+), folds were partitioned such that the proportion of EC tau positivity from each cohort was preserved in each of the ten folds.

#### 2.7.2 Aim 1: Using longitudinal Tau PET to validate SILA

##### Aim 1a: Assessing consistency of SILA-modeled tau trajectories across folds using 10-fold cross-validation

For each cohort, region, and fold in the 10-fold cross-validation (“KF” schema), SILA was trained on 90% of longitudinal tau PET SUVR data, with the remaining 10% used for testing—ensuring no overlap across test sets. Separately, SILA was also trained on the full dataset for each cohort and region (“WH” schema). Given the sparsity of longitudinal tau PET data, using all available data for model training may improve efficiency and facilitate application, provided it remains unbiased. For each cohort and region, the 10-fold-derived SUVR vs. estimated T+ time curves and the WH curve were plotted to compare SILA-estimated tau trajectories and identify model divergence. Additionally, participant-level longitudinal SUVR data were aligned to the modeled curves using the last scan as a reference point to visualize fit.

##### Aim 1b: SILA Tau PET SUVR and T+/- prediction accuracy

For all Aim 1b analyses, SILA models from Aim 1a (KF and WH schemas) were used to estimate SUVR and T+/– status across cohorts (ADNI, WISC) and ROIs. Four prediction methods were evaluated: (1) KF-backward, (2) KF-forward, (3) WH-backward, and (4) WH-forward. We investigated both forward and backward SILA prediction schemes to determine if, similar to our past results with amyloid PET, tau PET prediction accuracy was better for backwards vs. forward prediction (Betthauser et al., 2022).

Forward prediction used each participant’s first scan as the reference to estimate future SUVR and T+/– status, while backward prediction used the last scan to estimate earlier observed values. SUVR residuals (observed – predicted) were computed for each method and examined by sex, *APOE-e4* status, and cognitive status (non-dementia vs. dementia, defined as CDR global ≥1). Associations with age at reference scan, reference SUVR, and ETOA were also assessed.

Residuals were evaluated using residual sum of squares (RSS), bias, root mean square error (RMSE), and T+/– prediction accuracy. Because we expect a higher number of T-than T+, we also report balanced accuracy. Raw residual differences were first explored via Pearson correlations, then tested across categorical groups using Kruskal-Wallis tests. Linear mixed-effects models were used to assess associations with ETOA, reference SUVR, and age.

Sensitivity analyses, limited to the ADNI cohort, included using partial volume–corrected (PVC) data and restricting to non-demented participants (CDR <1). These were not performed in WISC due to few dementia cases (n = 2) and low atrophy, making PVC unnecessary.

##### Aim 1c: ETOA accuracy evaluation relative to observed T+ onset

For each cohort and ROI, SILA was used to estimate the age at which participants crossed the T+ threshold (estimated tau onset age; ETOA), using only the most accurate prediction methods validated in analyses 1a and 1b. ETOA accuracy was assessed in participants who converted from T– to T+ during follow-up: ADNI EC (n=28, 7.3%), ADNI MT (n=21, 5.5%), WISC EC (n=21, 7.3%), and WISC MT (n=18, 6.3%).

Observed T+ onset age was approximated as the midpoint between the last T– and first T+ scans. Midpoint conversion error was defined as the difference between this observed onset and the SILA-estimated ETOA. Accuracy was also reported as the proportion of ETOAs falling within the observed T– to T+ timeframe. Sensitivity analyses were conducted in ADNI using PVC data and excluding dementia cases.

#### 2.6.2 Aim 2: SILA-derived T+ rates, timing, and onset relate to common AD risk factors

##### Aim 2a: T+ rates in relation to APOE-e4 and dementia

To evaluate whether SILA-estimated tau positivity was more common among individuals with AD-associated factors, Fisher’s Exact Tests compared T+ proportions by *APOE*-e4 status and cognitive status at last visit (dementia-free vs. dementia). Analyses considered lifetime tau positivity.

The best performing SILA prediction methods from analyses 1a and 1b were used to assess T+ status. Odds ratios (ORs) were calculated for *APOE-e4* carriage and dementia presence using both threshold-based (i.e. observed) and SILA-indicated T+ to compare effect sizes and consistency between observed and estimated associations.

##### Aim 2b: T+ timing in relation to APOE-e4 and dementia

To assess ETOA differences by *APOE-e4* status, analyses were restricted to participants who were SILA-defined T+ in each ROI by the end of follow-up, including both converters (from 1c) and those entering the study T+. Kruskal-Wallis (KW) tests compared median ETOA by *APOE-e4* status, separately for EC and MT ROIs in ADNI and WISC, using each validated prediction method (from 1a and 1b). Results were stratified by prediction method. Sensitivity analyses were conducted in PVC data and the dementia-free subset in ADNI.

To assess differences in ETOA by *APOE-e4* status while controlling for baseline age, F-tests were conducted using linear models with ETOA as the response variable and *APOE-e4* status and baseline age as covariates. This analysis aimed to determine whether participants with different *APOE-e4* statuses show differences in ETOA, even when enrolling in the studies at approximately the same age.

To test whether individuals with dementia had spent more time T+ at their reference scan than dementia-free participants, KW tests compared median time T+ (years from ETOA to reference scan age) by cognitive status. Analyses were conducted separately by ROI, cohort, and prediction method, with sensitivity analyses in ADNI PVC data.

All tests were two-tailed with significance of α = 0.05.

## 3. RESULTS

### 3.1 Summary of study populations

Demographic characteristics for ADNI and WISC are shown in Table 1. On average, ADNI participants were older at first tau PET scan (mean difference: +6.0 years [4.95, 7.08], *p* < 0.001), had fewer females (*p* < 0.001), and more non-white individuals (*p* = 0.04) than WISC. ADNI had higher rates of CDR = 0.5 (32.7% vs. 12.5%) and dementia (i.e., CDR ≥1; 14.3% vs. 0.7%) at the last tau PET scan (both *p* < 0.001). The cohorts did not differ by *APOE* genotypes or *APOE-e4* carriage rates (45.5% vs. 40.6%, *p* = 0.16).

**Table 1.**
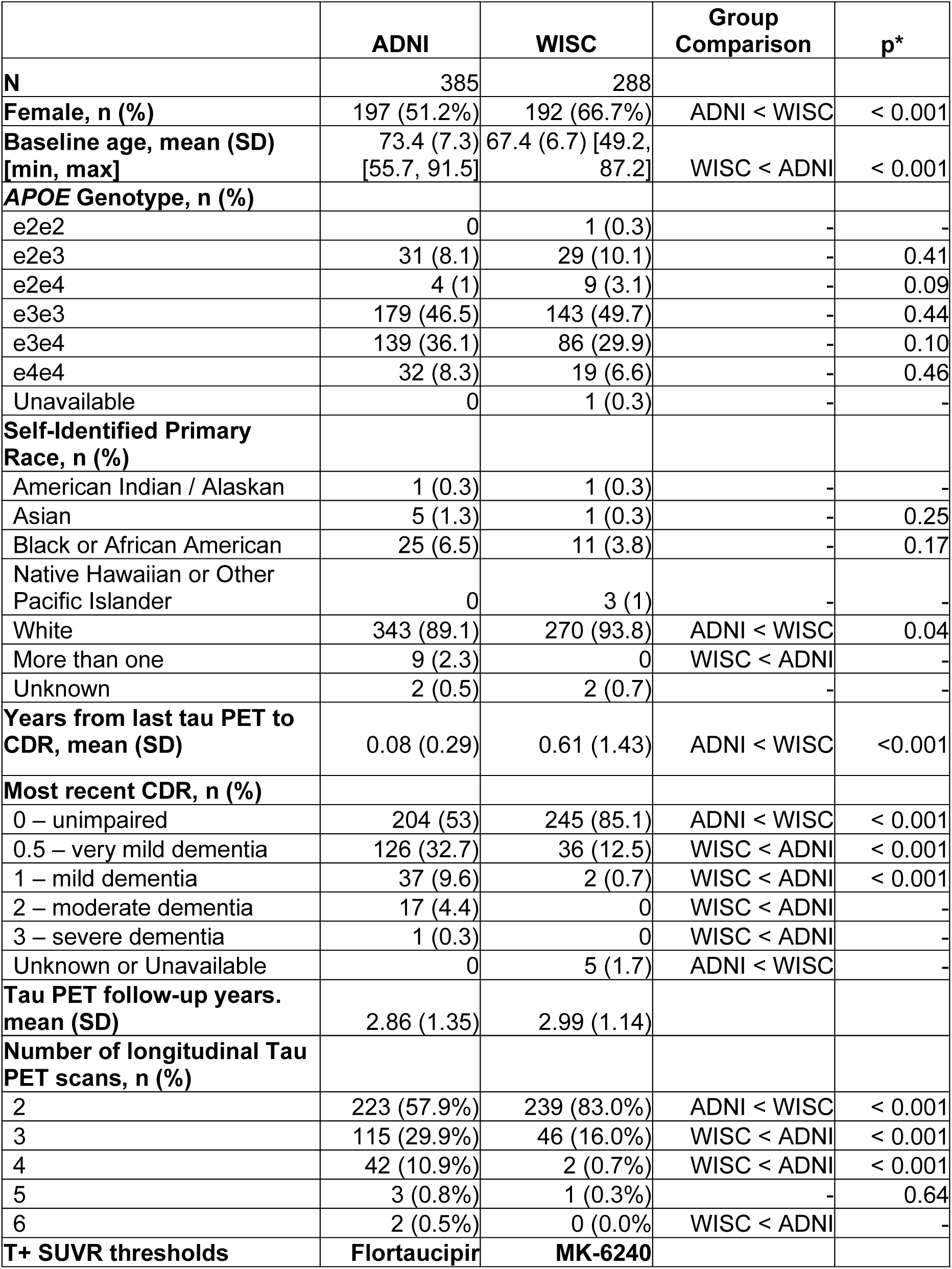
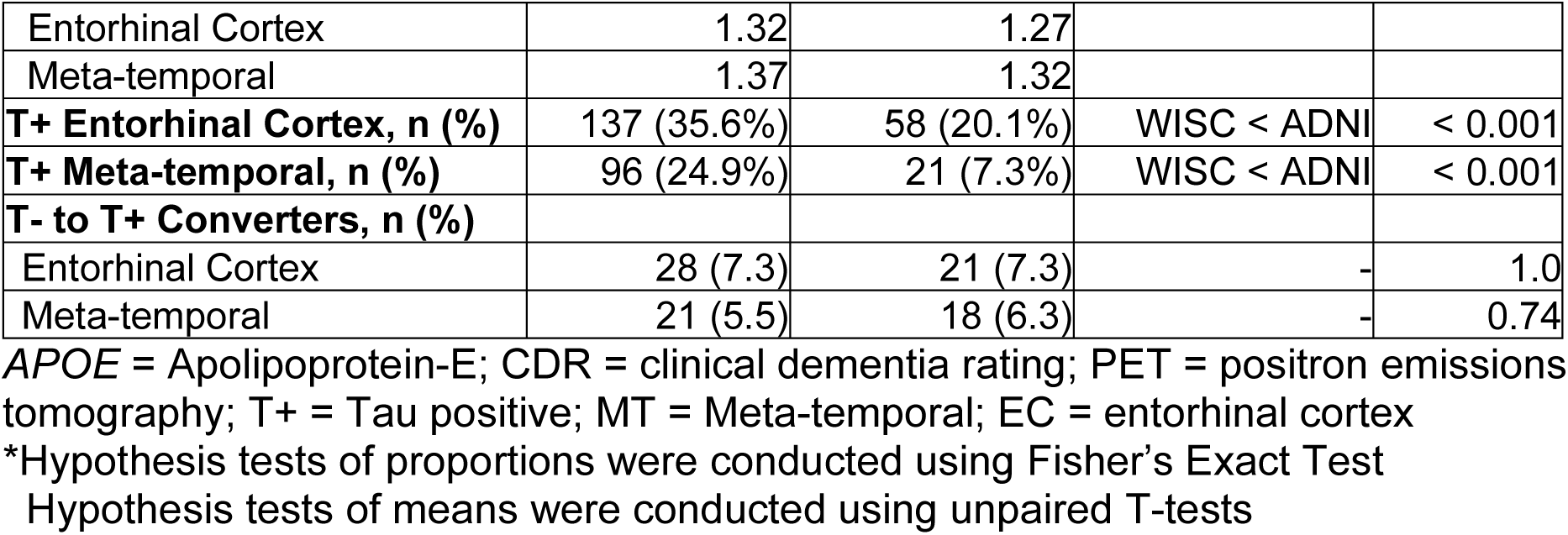
Cohort Characteristics.

Using cohort- and region-specific GMM thresholds, ADNI had higher T+ rates in both EC (35.6% vs. 20.1%) and MT ROIs (24.9% vs. 7.3%) (*p* < 0.001 for both). Mean follow-up was similar (ADNI: 2.87 [1.35] years; WISC: 2.99 [1.14] years, *p* = 0.22), with both cohorts having a median of two scans and maximum follow-ups of 5.88 years (6 scans, ADNI) and 4.90 years (5 scans, WISC). T– to T+ conversion rates did not differ significantly by cohort in either ROI (EC *p* = 1.0; MT *p* = 0.74).

### 3.2 Aim 1: Validity evidence for SILA prediction performance

On average, SILA model training and participant-level estimation took mean (SD) 14.90 (0.46) seconds for ADNI datasets and 10.46 (0.01) seconds for WISC datasets.

#### 3.2.1: Aim 1a) Examination of SILA-aligned trajectories

Figure 2 shows observed tau PET SUVR vs. age, SILA-modeled SUVR vs. estimated T+ time across folds, and participant-level, time-aligned SUVR vs. T+ time by cohort and region. Across all cohorts, ROIs, and folds, SILA produced monotonically increasing curves, with low slopes below the T+ threshold and steeper slopes above. Inflection points generally occurred just below the T+ threshold. MT SUVR curves were consistent across folds, while EC curves showed slight variability at higher SUVR values, likely due to sparse data and negative within-person slopes. Excluding dementia cases (CDR ≥1) in ADNI improved fold consistency for EC (Figure 3, Supplemental Figure 1). Participant-level alignment to the modeled curves was generally good, except for some T+ individuals with declining SUVR, primarily in the EC.

**Figure 2:**
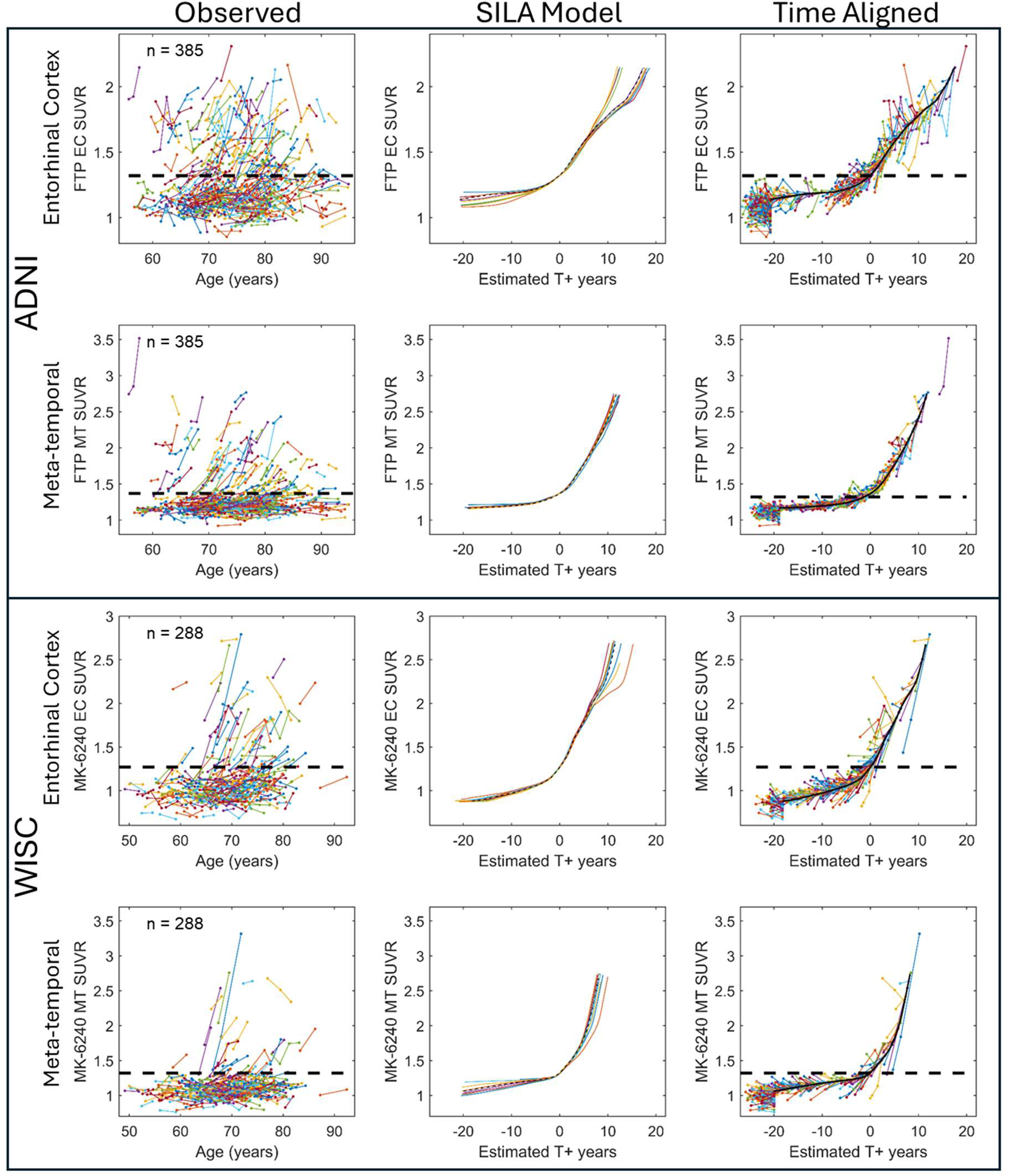
Observed and SILA-modeled Tau PET SUVR. Observed (left column), SILA-modeled (center column), and participant time-aligned (right column) tau PET SUVR curves. Different colors in columns 1 and 3 represent individual participants with horizontal dashed lines indicating region and cohort specific T+ SUVR thresholds. Colored lines in column 2 represent SILA-modeled tau PET SUVR vs. T+ time curves for each of the ten folds with the dashed black line indicating the modeled curve trained on all the data. EC = entorhinal cortex, MT = meta-temporal, FTP = Flortaucipir, SUVR = standard uptake value ratios, SILA = sampled iterative local approximation, PET = positron emission tomography.

**Figure 3:**
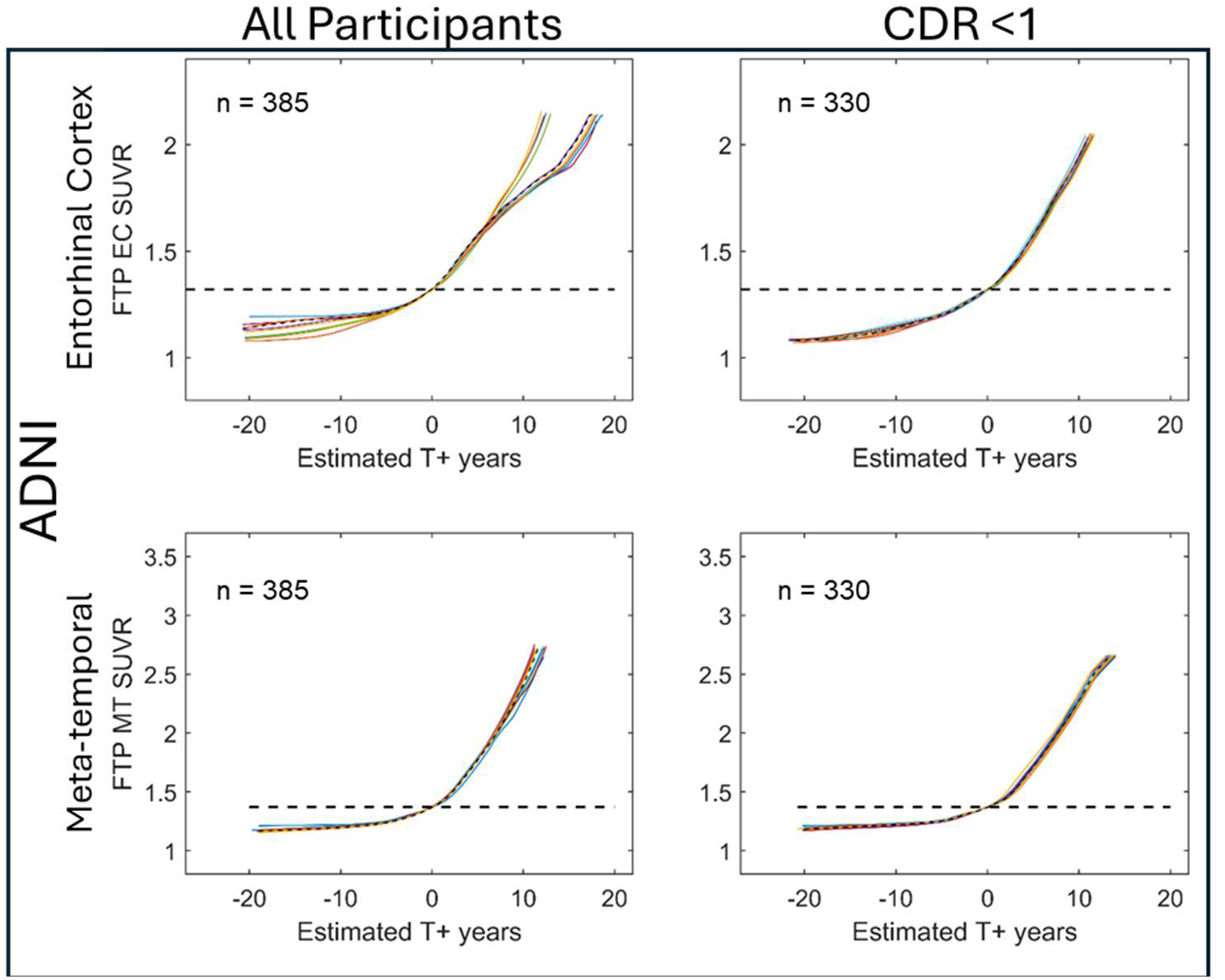
SILA model converge with and without dementia cases included (ADNI) SILA-modeled tau PET SUVR vs. T+ time curves in ADNI for all participants (left) and excluding participants with dementia (CDR ≥1; right). Each colored line represents the modeled relationship between SUVR and T+ time for each of ten folds with the black line representing the modeled relationship using all available data. Model convergence at higher SUVR values improved in entorhinal cortex when omitting participants with dementia from the dataset.

#### 3.2.2: Aim 1b) Forward and backward prediction of tau PET SUVR and T+/-

##### Model Performance Overview

Tables 2 and 3 summarize SUVR and T+/– prediction performance for ADNI and WISC across EC and MT ROIs using forward and backward methods. Backward prediction consistently outperformed forward prediction across all cohort/ROI combinations, with lower RMSE and RSS, comparable or lower bias (except ADNI MT), and higher T+/–accuracy (90–97% vs. 88–92%). MT ROIs similarly showed better overall performance (lower SUVR RSS, RMSE, bias; higher T+/- accuracy) than EC.

**Table 2.** Ten-fold cross-validation SILA model summary statistics.

| Prediction Schema | Metric | ADNI |  | WISC |  |
| --- | --- | --- | --- | --- | --- |
|  |  | Entorhinal | Meta-temporal | Entorhinal | Meta-temporal |
| **Backward** | **RSS SUVR** | 23.21 | 17.41 | 25.9 | 20.8 |
|  | **RMSE SUVR** | 0.065 | 0.049 | 0.090 | 0.072 |
|  | RMSE(T+) | 0.11 | 0.12 | 0.232 | 0.247 |
|  | RMSE(T-) | 0.05 | 0.03 | 0.071 | 0.060 |
|  | **Bias SUVR** | 0.022 | 0.021 | 0.042 | 0.019 |
|  | **T+/-**<br>**Balanced**<br>**Accuracy** | 84.8% | 90.4% | 81.1% | 88.3% |
|  | **T+/-**<br>**Accuracy** | 87.6% | 96.1% | 93.8% | 97.6% |
|  | **Number of T-**<br>**to T+**<br>**converters** | 28 | 21 | 21 | 18 |
|  | **Conversion**<br>**accuracy, %**<br>**(n<sub>corr</sub>)** | 75.0%<br>(21/28) | 76.2%<br>(16/21) | 85.7%<br>(18/21) | 77.8%<br>(14/18) |
|  | **Conversion**<br>**midpoint**<br>**error mean**<br>**years [95%**<br>**CI]** | -0.33<br>[-0.78, 0.13] | 0.12<br>[-0.27,<br>0.52] | -0.09<br>[-0.64, 0.46] | -0.09<br>[-0.93,<br>0.74] |
| **Forward** | **RSS SUVR** | 28.2 | 25.6 | 41.0 | 28.4 |
|  | **RMSE** | 0.081 | 0.072 | 0.142 | 0.098 |
|  | RMSE(T+) | 0.120 | 0.136 | 0.271 | 0.271 |
|  | RMSE(T-) | 0.065 | 0.053 | 0.119 | 0.119 |
|  | **Bias SUVR** | 0.042 | 0.019 | -0.075 | -0.030 |
|  | **T+/-**<br>**Balanced**<br>**Accuracy** | 85.2% | 92.6% | 78.8% | 84.7% |
|  | **T+/-**<br>**Accuracy** | 83.3% | 91.2% | 78.2% | 91.0% |

Model summary statistics for backward and forward SUVR and T+/- prediction. Backwards prediction was used to validate estimated T+ onset age (ETOA) in a subset of observed T- to T+ converters (gray) from 10-fold cross-validation results. Conversion accuracy was defined as the number of times ETOA was between the observed last T- and first T+ scan for observed converters. Conversion midpoint error was defined as the difference between ETOA and the midpoint between the last T- and T+ scan for observed converters. RMSE = root mean squared error, RSS = residual sum of squares, SUVR = standard uptake value ratio

**Table 3.**
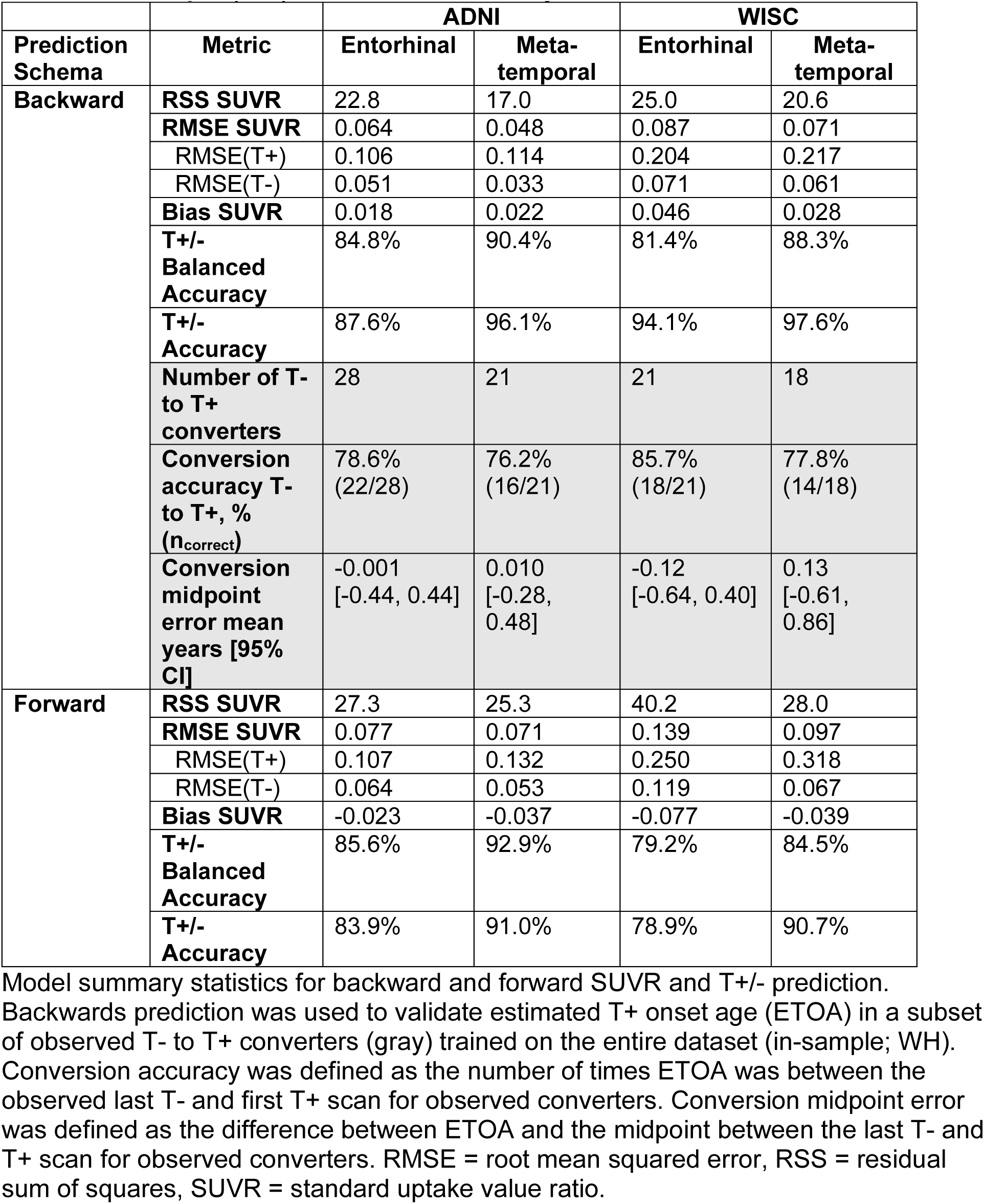
In-sample (WH) SILA model summary statistics.

Whole-sample (WH) training slightly outperformed 10-fold (KF) training, with backward model bias around –0.02 to –0.01 SUVR (vs. 0.01 to 0.04 SUVR for forward). WH-backward RMSE (0.09 SUVR) was marginally lower than KF-backward (0.10 SUVR), suggesting a slight accuracy gain—likely due to overfitting—but without substantial bias relative to KF. Excluding dementia cases in ADNI improved performance across all methods (Supplemental Table S1). However, using partial volume–corrected (PVC) data worsened SUVR prediction across all metrics (Supplemental Table S2).

##### SUVR Residual Analysis

Figures 4 and 5 illustrate residuals by reference SUVR, time from reference scan, and age in the MT ROI and EC, respectively. While no consistent patterns emerged, higher residual variability was observed in T+ participants (high reference SUVR) and, in WISC, in older individuals and those with longer intervals from the reference scan—likely due to data sparsity. Backward and forward prediction residuals showed low to moderate correlations (–0.27 to 0.01) with reference SUVR and ETOA in both ADNI and WISC (Supplemental Tables S3-S10). Residual correlations with sex were minimal (|r| < 0.03) and were highest in ADNI PVC data for entorhinal cortex using KF-backward (r = 0.09).

**Figure 4:**
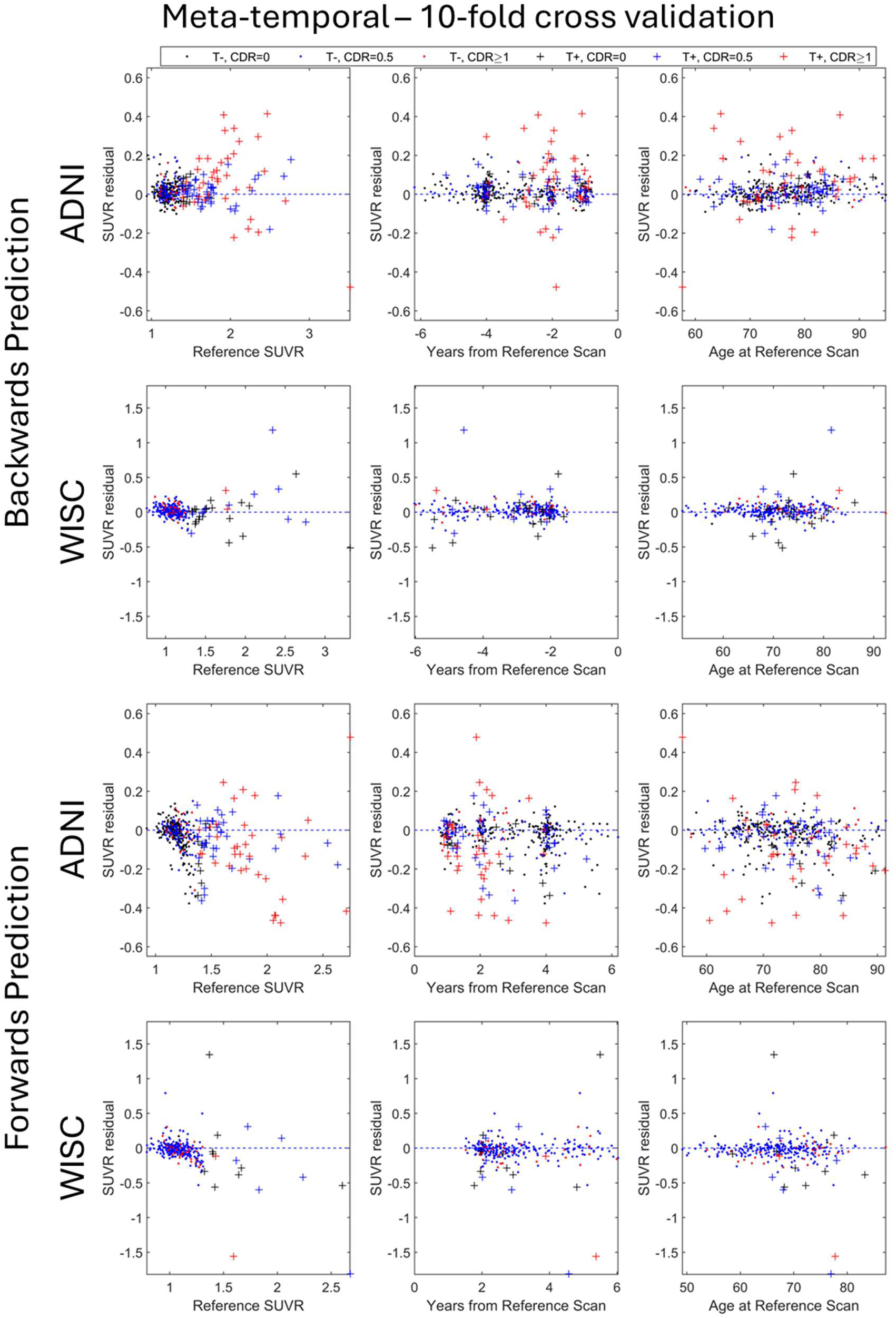
SILA meta-temporal SUVR prediction residuals. Backward (top two rows) and forward (bottom two rows) SILA prediction residuals (observed – estimated) in the meta-temporal ROI from 10-fold cross-validation as a function of age, reference SUVR, and time from reference scan. Color indicates CDR global score at the last available tau PET scan.

**Figure 5:**
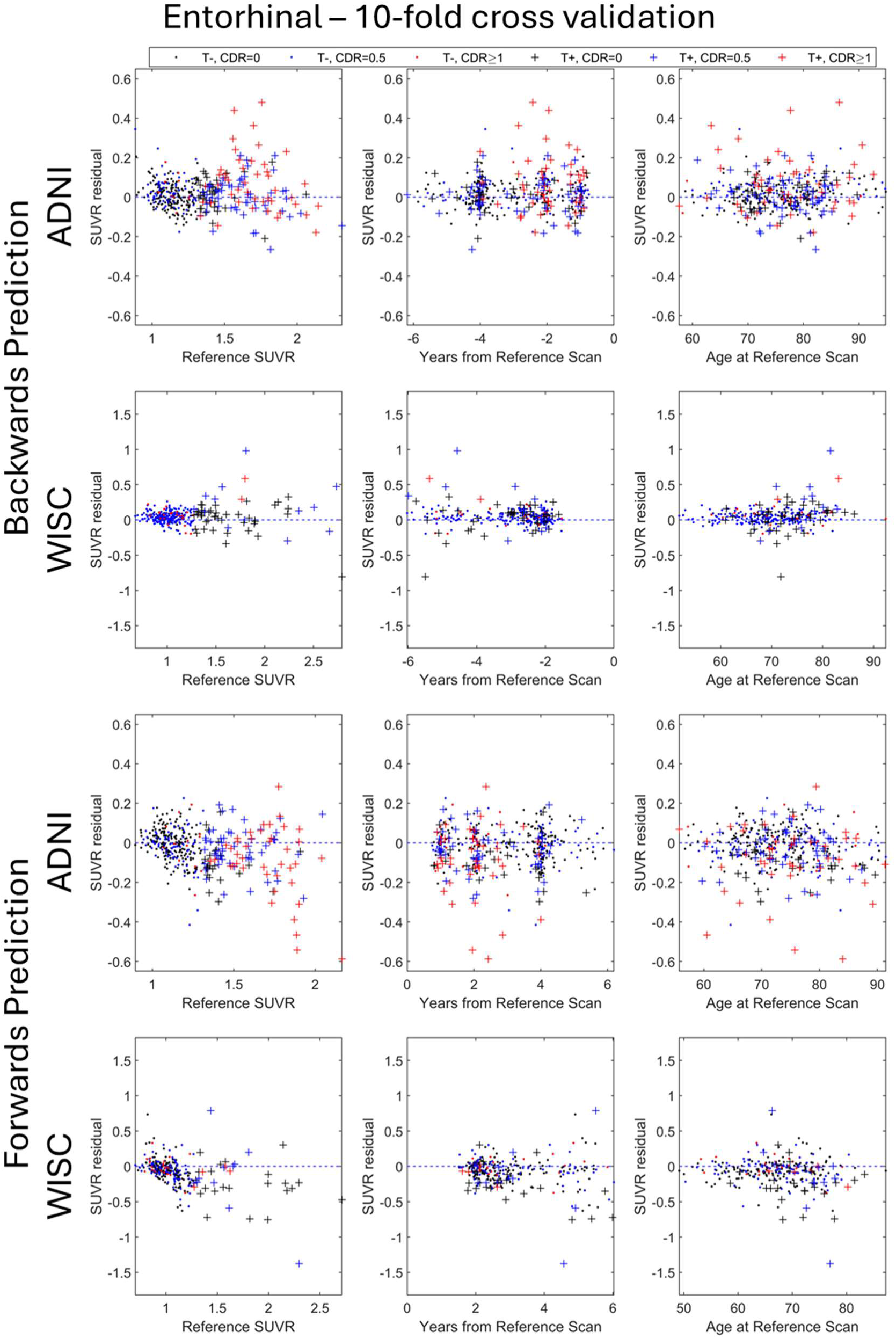
SILA Entorhinal Cortex SUVR prediction residuals. Backward (top two rows) and forward (bottom two rows) SILA prediction residuals (observed – estimated) in the entorhinal cortex ROI from 10-fold cross-validation as a function of age, reference SUVR, and time from reference scan. Color indicates CDR global score at the last available tau PET scan.

Kruskal-Wallis tests revealed no statistically significant difference between *APOE-e4*+ and *APOE-e4*- individuals in backward prediction residuals for either cohort or ROI, however, WH forward prediction did show statistically significant differences mainly in the MT ROI (overestimated for e4- individuals 0.007 SUVR more than e4+ individuals; p = 0.04). This was consistent in the dementia-free data for the ADNI MT ROI using both forward prediction methods (overestimated for e4- individuals 0.012 SUVR more than e4+ individuals in both prediction methods; p_WH_ = 0.001 and p_KF_ = 0.002).

Between those with dementia and those dementia-free in ADNI, forward prediction in the EC ROI overpredicted tau SUVR for those with dementia to a greater degree than those dementia-free (SUVR Diff_WH_ = 0.03, p_WH_ = 0.02; Diff_KF_ = 0.03, p_KF_ = 0.05). All prediction methods had statistically significant differences between dementia and dementia-free SUVR prediction residuals. Backward prediction methods tended to underpredict SUVR in those with dementia more than those dementia-free (SUVR Diff_WH_ = 0.02, p_WH_ = 0.05; Diff_KF_ = 0.01, p_KF_ = 0.02). Forward prediction methods behaved similarly as in the EC ROI, with SUVR being overpredicted to a greater degree in those with dementia (SUVR Diff = 0.03 for both; p_WH_ = 0.02, p_KF_ = 0.002).

In ADNI, forward model residuals in the EC how low negative correlation with reference SUVR (r_KF_ = –0.24, r_WH_ = –0.17) and low positive correlation with ETOA (r_KF_ = 0.19, r_WH_ = 0.13). Similar but stronger trends were observed in the MT ROI (r_Ref_ = –0.28 to –0.25; r_ETOA_ = 0.19–0.20), especially in dementia-free participants and PVC data. In WISC, EC forward residuals were moderately associated with reference SUVR (r = –0.33) and ETOA (r = 0.23–0.25); MT residuals showed stronger negative correlations with reference SUVR (r = –0.40) and weaker positive associations with ETOA (r = 0.13–0.18). No other variables showed |r| > 0.14.

3.2.4: Aim 1c) ETOA prediction accuracy

To estimate ETOA accuracy, we used SILA-estimated ETOAs from backward prediction methods (KF and WH), which showed better performance in analysis (1b) and appropriate convergence in (1a) compared to forward prediction. These methods were applied to subsets of T– to T+ converters for each cohort and ROI.

Tables 2 and 3, Supplemental Tables S1 and S2, and shown in Figure 6 summarizes accuracy, midpoint conversion errors (MCEs), and value estimates for each prediction method by site/ROI. ETOA estimates fell between the last observed T– and first T+ scan in 72%–86% of cases, with the lowest accuracy in WISC MT (72%) and highest in WISC EC (86%). WH backward estimates generally had higher accuracy than KF, with WH predictions differing from midpoint ETOA by 0.69–1.22 years, compared to 0.71–1.38 years for KF. The mean differences between ETOA and the midpoint between the last T- and first T+ scan ranged from -0.33 to 0.12 years across ROIs and cohorts.

**Figure 6:**
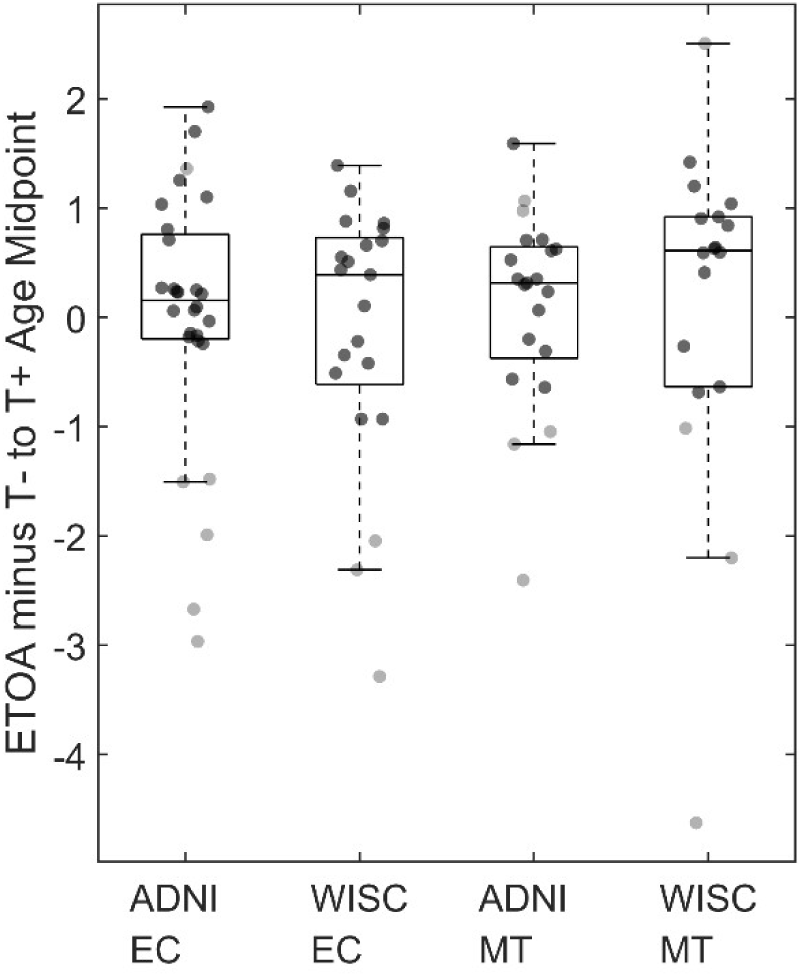
Accuracy of estimated Tau onset age (ETOA) in observed T- to T+ converters. Approximated error in the estimated T+ onset age from SILA by cohort and region using data derived from the SILA model trained on the entire dataset (WH). Error in SILA-estimated T+ onset age (ETOA) is reported as the difference in years between the SILA-estimated T+ onset age and the midpoint between the observed last T- scan and first T+ scan (midpoint) and. Solid points indicate that the SILA-estimated T+ age was within the interval between the last T- and first T+ scan with gray points indicating SILA estimates were outside of this range.

### 3.3 Aim 2: SILA-derived T+ rates, timing, and onset relate to common AD risk factors

#### 3.3.1: Aim 2a) T+ rates in relation to APOEe4+ and dementia

##### T+ rates between APOEe4+ and APOEe4-

Fisher’s Exact Test assessed whether SILA-defined tau positivity, using backward prediction, was more prevalent among *APOE-e4* carriers. Odds ratios (ORs) for SILA-defined positivity vs. observed positivity (GMM threshold) were compared to evaluate consistency in T+ cases (Table 4). Across all ROIs and cohorts, *APOE-e4* carriage was associated with increased odds of tau positivity by SILA. In ADNI, lifetime SILA-estimated tau positivity odds ranged from 3.86 (MT) to 4.16 (EC) times higher for *APOE-e4+* vs*. APOE-e4-* using KF-backward prediction, and 3.86 (MT) to 4.07 (EC) with WH-backward prediction. Observed ORs were 3.78 [2.76, 5.22] (MT) and 4.22 [3.17, 5.63] (EC) for *APOE-e4*+. KF-backward prediction ORs were closer to observed ORs, maintaining the directionality of increased odds in *APOE-e4+*. This result held when excluding dementia cases in ADNI (Table 4). In PVC data, directionality was preserved, with WH-backward ORs closer to observed ORs (WH differences = 0.00 in MT, 0.01 in EC; KF differences = 0.11 in MT, 0.01 in EC; Supplemental Table S12).

**Table 4.** Lifetime T+ Odds Ratios for *APOE-e4* carriers.

| Entire Dataset | ADNI EC | ADNI MT | WISC EC | WISC MT |
| --- | --- | --- | --- | --- |
| Observed T+ | 4.22<br>(3.17, 5.63)* | 3.78<br>(2.76, 5.22)* | 3.20<br>(2.11, 4.89)* | 4.43<br>(2.59, 7.79)* |
| Estimated T+ (KF) | 4.16<br>(2.62, 6.70)* | 3.86<br>(2.30, 6.60)* | 3.46<br>(1.83, 6.69)* | 5.09<br>(2.18, 12.95)* |
| Estimated T+ (WH) | 4.07<br>(2.56, 6.55)* | 3.86<br>(2.30, 6.60)* | 3.46<br>(1.83, 6.69)* | 5.76<br>(2.39, 15.40)* |
| **PVC** |  |  |  |  |
| Observed T+ | 4.22<br>(3.18, 5.63)* | 3.78<br>(2.76, 5.22)* | - | - |
| Estimated T+ (KF) | 3.87<br>(2.45, 6.19)* | 3.61<br>(2.19, 6.04)* | - | - |
| Estimated T+ (WH) | 3.87<br>(2.45, 6.19)* | 3.36<br>(2.05, 5.59)* | - | - |
| **Dementia-free** |  |  |  |  |
| Observed T+ | 4.03<br>(2.92, 5.60)* | 3.25<br>(2.18, 4.91)* | - | - |
| Estimated T+ (KF) | 3.76<br>(2.21, 6.49)* | 3.52<br>(1.87, 6.82)* | - | - |
| Estimated T+ (WH) | 3.76<br>(2.21, 6.49)* | 3.52<br>(1.87, 6.82)* | - | - |

Lifetime T+ risk reported as odds ratios with 95% confidence intervals
\* indicates p<0.05
EC = entorhinal cortex, KF = k-fold results, MT = meta-temporal, PVC = partial volume corrected, WH = whole cohort results

In WISC, ORs for lifetime tau positivity between *APOE-e4* carriers and non-carriers ranged from 4.12 (EC) to 4.94 (MT) for KF-backward prediction, and 4.21 (EC) to 5.52 (MT) for WH-backward prediction. Observed ORs were 5.68 [2.97, 11.48] (MT) and 3.50 [2.21, 5.59] (EC) for *APOE-e4+*. KF-backward prediction yielded ORs closer to observed values while maintaining the directionality of increased odds for *APOE-e4*+. Differences between observed and estimated ORs were greater in WISC than in ADNI and remained within 95% confidence intervals.

##### T+ rates and dementia

Due to few dementia cases in WISC (n=2), dementia effects were assessed only in ADNI. Fisher’s test showed a significant increase in tau positivity for those diagnosed with dementia by their last visit. Lifetime KF-backward ORs [95% CI] ranged from 10.39 [5.31, 21.09] in the MT ROI to 11.73 [5.54, 27.26] in the EC ROI, while WH-backward ORs ranged from 10.39 [5.31, 21.09] in the MT ROI to 11.91 [5.62, 27.67] in the EC ROI (Table 5). PVC-based sensitivity analyses were consistent with these results. Observed ORs [95% CI] were 11.89 [7.34, 20.00] in the EC ROI and 10.56 [6.92, 16.32] in the MT ROI. While KF-backward prediction was closer to observed ORs, wide confidence intervals suggest the exact OR values are less precise than for *APOE-e4+* individuals.

**Table 5.** Lifetime T+ Odds Ratios for Participants with Dementia.

| Entire Dataset | ADNI EC | ADNI MT |
| --- | --- | --- |
| Observed T+ | 11.89 (7.34, 20.0) * | 10.56 (6.92, 16.32) * |
| Estimated KF T+ | 11.73 (5.54, 27.26) * | 10.39 (5.31, 21.09) * |
| Estimated WH T+ | 11.91 (5.62, 27.67) * | 10.39 (5.31, 21.09) * |
| **PVC** |  |  |
| Observed T+ | 12.39 (7.59, 21.05) * | 10.80 (7.06, 16.75) * |
| Estimated KF T+ | 9.60 (4.64, 21.52) * | 10.91 (5.50, 22.65) * |
| Estimated WH T+ | 9.60 (4.64, 21.52) * | 10.50 (5.30, 21.77) * |

Lifetime T+ risk reported as odds ratios with 95% confidence intervals
\* indicates p<0.05
EC = entorhinal cortex, KF = k-fold results, MT = meta-temporal, PVC = partial volume corrected, WH = whole cohort results

#### 3.3.2: Aim 2b) T+ timing in relation to *APOE-e4*+ and dementia

##### Estimated Tau Onset Age vs. APOE-e4+

Data were subset to include only those who were T+ by the end of observation separately for each ROI. KW tests examined differences in ETOA distribution by *APOE-e4* positivity. In ADNI’s EC, ETOAs were significantly earlier in *APOE-e4*+ individuals across all prediction methods (*p* < 0.01), with *APOE-e4*+ showing onset 4.6 (KF) - 4.7 (WH) years earlier than *APOE-e4*- (Table 6). This difference was also significant in the MT region in ADNI, with *APOE-e4*+ associated with ETOAs 3.8 years earlier than *APOE-e4*- across both prediction methods. These results were consistent using PVC data. For dementia-free cases (ADNI) these results were consistent in the EC ROI but did not reach significance in the MT ROI using either prediction method (p(KF) = 0.11, p(WH) = 0.12).

**Table 6:** ETOA by *APOE-e4* carriage.

| Entire Dataset | ADNI EC | ADNI MT | WISC EC | WISC MT |
| --- | --- | --- | --- | --- |
| KF-backward | 4.7 (< 0.001) | 3.8 (0.004) | 4.7 (< 0.001) | 2.1 (0.34) |
| WH-backward | 4.6 (< 0.001) | 3.8 (0.003) | 4.5 (< 0.001) | 2.9 (0.24) |
| **PVC** |  |  |  |  |
| KF-backward | 4.3 (< 0.001) | 3.9 (0.002) | - | - |
| WH-backward | 4.4 (< 0.001) | 3.9 (0.002) | - | - |
| **Dementia-free** |  |  |  |  |
| KF-backward | 3.5 (0.001) | 1.8 (0.11) | - | - |
| WH-backward | 3.7 (0.001) | 1.9 (0.12) | - | - |

Median absolute difference in ETOA between *APOE-e4* non-carriers minus carriers (KW Test p-value)
EC = entorhinal cortex, KF = k-fold results, MT = meta-temporal, PVC = partial volume corrected, WH = whole cohort results

In WISC, EC ETOAs were significantly earlier in *APOE-e4*+ participants across all prediction methods (*p* ≤ 0.01), with differences ranging from 4.5 (WH) to 4.7 (KF) years earlier in *APOE-e4*+ individuals. In the MT ROI, differences in ETOAs were earlier for *APOE-e4+* but failed to be significant in both backward prediction methods (Table 6; p(KF) = 0.34, p(WH) = 0.24).

Overall, results suggest *APOE-e4* carriage is consistently associated with earlier tau onset as indicated by SILA in the EC, with estimated onset occurring 4.6 years earlier than *APOE-e4*-, on average, among those observed to become T+. This pattern was less consistent in the MT ROI across cohorts, potentially due to regional variation in tau accumulation and differences in cohort composition. Using linear models controlling for baseline tau PET age, *APOE-e4* carriage remained a significant predictor of ETOA in the EC, whereas *APOE-e4* carriage was no longer significantly associated with MT ETOA in ADNI (*p* range 0.15 – 0.40) with mixed results in WISC (*p* range 0.02 – 0.06) (Table 7).

**Table 7:** ETOA by *APOE-e4* carriage controlling for baseline age.

| Entire Dataset | ADNI EC | ADNI MT | WISC EC | WISC MT |
| --- | --- | --- | --- | --- |
| KF-backward | -2.54<br>(-4.19, -0.88) | -1.14<br>(-2.74, 0.44) | -2.84<br>(-4.81, -0.88) | -2.50<br>(-5.12, 0.13) |
| WH-backward | -2.63<br>(-4.37, -0.90) | -1.13<br>(-2.67, 0.40) | -2.79<br>(-4.63, -0.95) | -3.58<br>(-6.50, -0.66) |
| **PVC** |  |  |  |  |
| KF-backward | -2.59<br>(-4.82, -0.36) | -1.39 (-3.14, 0.36) | - | - |
| WH-backward | -2.64<br>(-4.44, -0.84) | -1.35<br>(-3.05, 0.35) | - | - |
| **Dementia-free** |  |  |  |  |
| KF-backward | -1.58<br>(-3.01, -0.15) | -0.61<br>(-2.49, 1.28) | - | - |
| WH-backward | -1.65<br>(-3.08, -0.22) | -0.68<br>(-2.61, 1.25) | - | - |

$\beta(e4+)$ (95% CI)
EC = entorhinal cortex, KF = k-fold results, MT = meta-temporal, PVC = partial volume corrected, WH = whole cohort results

##### Time T+ and Dementia

KW-tests assessed whether individuals with dementia had been T+ longer at reference scan (final scan). In ADNI, dementia was associated with significantly longer SILA-estimated T+ time in the EC across all prediction methods (p < 0.001), with estimates ranging from 4.0 (KF) - 4.1 (WH) years longer in those without dementia at last visit (Table 8). This was also the case in the MT region, with dementia associated with longer T+ time of 3.2 years by last visit for both prediction methods. These results were consistent in the PVC data. We did not explore this relationship in the WISC data due to low dementia presence (n = 2).

**Table 8:** Estimated T+ time by dementia status.

| Entire Dataset | ADNI EC | ADNI MT |
| --- | --- | --- |
| KF-backward | 4.04 (< 0.001) | 3.20 (< 0.001) |
| WH-backward | 4.10 (< 0.001) | 3.20 (< 0.001) |
| **PVC** |  |  |
| KF-backward | 6.03 (< 0.001) | 3.46 (< 0.001) |
| WH-backward | 4.78 (< 0.001) | 3.46 (< 0.001) |

Difference in SILA-estimates T+ time between those with dementia minus those without dementia (p-value)
EC = entorhinal cortex, KF = k-fold results, MT = meta-temporal, PVC = partial volume corrected, WH = whole cohort results

## 4. Discussion

The overarching goal of this work was to determine the validity of SILA for modeling longitudinal tau PET trajectories and estimating individual T+ onset ages in two different cohorts using different tau PET tracers. Our results indicate that SILA accurately estimates SUVR and T+ onset ages retrospectively in the MT ROI regardless of age, *APOE-e4*, and dementia status whereas SILA time and SUVR estimates in EC were impacted by the presence of dementia. Similar to amyloid PET, retrospective and prospective estimation perform reasonably well in those that are T+, whereas retrospective estimation significantly outperforms prospective estimation among T-individuals. Utilizing the entire data sample as opposed to a classical train/test split (KF) resulted in similar estimates, with minimal impact on model bias. Lastly, SILA-estimated values were associated with established dementia risk factors and did not dramatically alter relationships between established dementia risk factors compared to observed tau PET outcomes. Taken together, these results suggest SILA can be used to characterize longitudinal tau PET trajectories, estimate individual T+ time and age, and offer insights into longitudinal tau PET changes and the predictability of tau accumulation measured with tau PET imaging.

Similar to prior studies modeling longitudinal amyloid PET trajectories (Betthauser et al., 2022; Bilgel et al., 2016; Cogswell et al., 2024; Koscik et al., 2020; Schindler et al., 2021), our results suggest tau PET accumulation appears to be highly predictable once above PET detection limits and it is likely that several methods (Therneau et al., 2021; Whittington et al., 2021) can be used to accurately characterize the longitudinal time course of tau PET accumulation. Similarly, common AD risk factors like age, sex, and *APOE-e4,* while associated with T+ risk and age, do not appear to greatly influence this overall accumulation trajectory. This observation suggests these factors influence *when* pathology begins to accumulate, but once pathological tau aggregation begins within a brain region, it will continue to accumulate at a consistent and predictable rate. One caveat is that clinical status (i.e., the presence of dementia) and brain region appear to more strongly influence longitudinal tau PET SUVR compared to amyloid PET (Betthauser et al., 2022). For example, our analyses showed that entorhinal cortex SUVR residuals were associated with dementia status, and that model convergence and fit improved in the entorhinal cortex when omitting participants with CDR ≥1, whereas model performance was similar when including or excluding CDR ≥1 and associations between dementia and model residuals were weaker in the meta-temporal ROI. As a result, the optimal methods for tau PET temporal modeling likely differ depending on the study population and study goals. Coupling our results with prior studies demonstrating the entorhinal cortex is typically the first region to show elevated tau (Braak & Braak, 1991; Cody et al., 2024; Pascoal et al., 2021) suggests that entorhinal cortex SUVR produces accurate tau-based time estimates during preclinical AD, but that the accuracy of these estimates diminishes as individuals accumulate higher levels of tau and transition to dementia. In contrast, composites such as the Jack meta-temporal used in our analysis (Jack et al., 2017), or universal CenTauR (Villemagne et al., 2023) ROIs likely sacrifice some early detectability but provide accurate tau-based time estimates spanning preclinical and clinical disease phases.

The worsening of SILA performance in entorhinal cortex with dementia appeared to be predominantly driven by participants with dementia being more likely to have negative within-person entorhinal cortex SUVR slopes at higher SUVR values (Figure 7). One possible explanation for this effect is that participants with dementia are expected to have greater medial temporal volume loss and would therefore exhibit higher partial volume effects on tau PET imaging. While partial volume correction weakened the association between SILA SUVR residuals and dementia, it also resulted in a near doubling of RMSE and introduced negative SUVR change in individuals wherein volume changes were slightly positive over time. This is consistent with a prior study suggesting that applying partial volume correction to tau PET data strengthens associations between tau PET SUVR and longitudinal atrophy, but only marginally impacts group differences in longitudinal tau PET change and therefore may not be beneficial in practice (Costoya-Sanchez et al., 2024). Another possible explanation for negative within-person entorhinal cortex slopes in participants with dementia is that tau tracer binding and access to tau binding sites may be affected by tau maturation and the transition from intraneuronal to extracellular tau tangles (Malarte et al., 2024), which is known to affect binding affinity of histochemical and immunohistochemical staining (Moloney et al., 2021). Regardless of mechanism, the higher frequency of negative within-person tau SUVR slopes needs to be considered when interpreting longitudinal change, particularly in the context of clinical trials wherein an observed lower SUVR over time could be incorrectly attributed to a treatment effect depending on the brain region analyzed.

**Figure 7:**
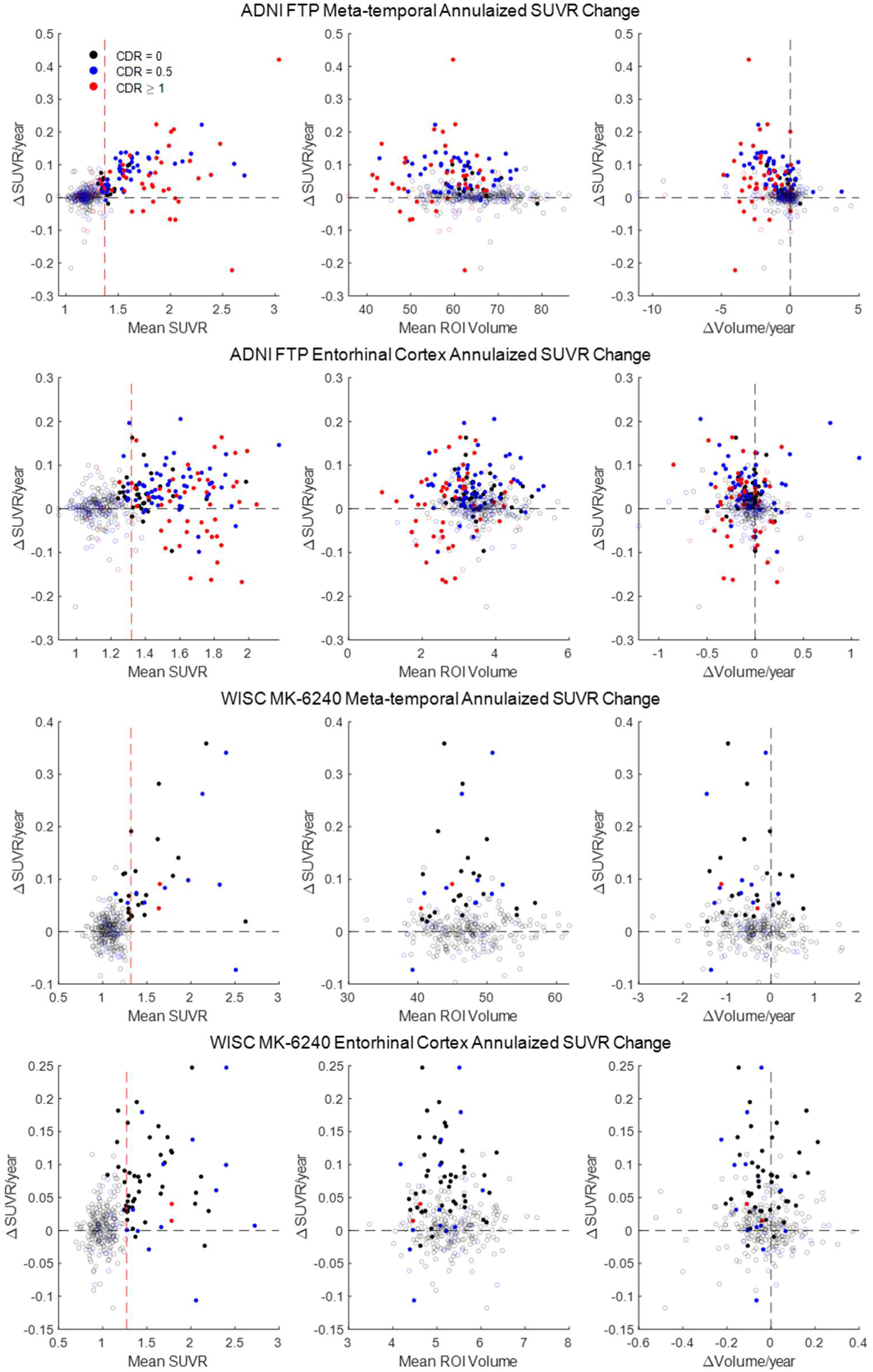
Rate of tau change by SUVR, ROI volume, and ROI annualized volume change. Observed within-person mean annualized tau PET SUVR change as a function of mean SUVR (left), mean ROI volume (middle), and within-person annualized volume change (right) for each cohort and ROI. Open circles represent individuals that are T- whereas closed circles are T+ at their last observation.

Our results suggest some best practices when implementing SILA and other approaches in longitudinal tau PET data. First, SILA performs best in a retrospective design, that is, estimating antecedent SUVR or T+ age rather than for prospective prediction of tau SUVR or when tau positivity will occur in the future. This is especially evident in T- individuals with SUVR values near but below the T+ threshold. In this range, SILA produces accurate estimates of antecedent SUVR values but overestimates future SUVR values for people that remain T- at later timepoints. Similar to SUVR values, the confidence that binding estimates in this range are due to tau and thereby that time estimates are reliable in this subthreshold range decrease as SUVR values decrease below PET detection limits. Regarding training schemes, in-sample training schemes did not greatly impact model estimates and may be preferred in sparse datasets, but given convergence issues related to dementia participants, we suggest verifying SILA convergence with an out-of-sample scheme like k-fold prior to implementing more broadly in additional datasets and brain regions not tested in this work. Lastly, SILA is not designed as a predictive algorithm to determine when people will become T+ in the future. While including additional factors associated with T+ risk like *APOE-e4*, sex, or other biomarkers like amyloid PET or plasma pTau217 would likely improve prediction of T+ age amongst T- people, this is not the design goal of the algorithm. By being able to retrospectively estimate T+ age, SILA and other similar approaches are well-suited for research applications identifying factors that contribute to this heterogeneity in T+ ages and the timing from T+ to clinical impairment and other pertinent disease events, particularly for people that are already T+ at their first observation and the transition from T- to T+ cannot be directly observed.

This work has a number of strengths and limitations to acknowledge. Among the strengths is the replication of results in two different cohorts using two different tau PET tracers in different brain regions. It is worth noting that analyses pertaining to the impact of dementia on SILA performance were limited to ADNI, since the WISC dataset only had two participants with dementia. Future work should continue to investigate the impact of dementia on regional longitudinal tau PET trajectories in additional cohorts. In addition, while both ADNI and WISC are actively making efforts to include underrepresented groups, the subsets with available longitudinal tau PET imaging analyzed here do not reflect the general population and therefore may not generalize across population groups.

In summary, our results provide strong evidence that SILA can accurately model longitudinal tau PET trajectories and provide estimates of T+ age and retrospective SUVR. In contrast to amyloid PET, additional considerations for disease stage and brain region need to be taken into account when assessing the accuracy of SILA-based time estimates derived from tau PET imaging.

## Data and Code Availability

Data used in the preparation of this article were obtained from the Alzheimer’s Disease Neuroimaging Initiative (ADNI) database (https://adni.loni.usc.edu), and WRAP and Wisconsin ADRC https://www.adrc.wisc.edu/apply-resources. Data from each study are available upon request and executed data transfer and use agreements as required by source studies. SILA code is available at the following github repository: https://github.com/Betthauser-Neuro-Lab/SILA-AD-Biomarker.

## Author Contributions

**TJ Betthauser:** conceptualization, methodology, software, formal analysis, writing-original draft, writing-review & editing, supervision, project administration, funding acquisition.

**JP Teague:** writing-original draft, writing-review & editing, formal analysis

**H Bruzzone:** formal analysis, manuscript writing-review & editing

**W Coath:** manuscript writing-review & editing

**MB Heston:** manuscript writing-review & editing

**JD Morse:** data curation, manuscript writing-review & editing

**EL Ruiz de Chavez:** data curation, manuscript writing-review & editing

**FJ Carey:** manuscript writing-review & editing, project administration

**R Navaratna:** manuscript writing-review & editing

**KA Cody:** manuscript writing-review & editing

**RE Langhough:** conceptualization, statistical methodology, data interpretation, manuscript writing-review & editing

## Data Availability

All data analyzed in this study are available upon online request to ADNI and WRAP cohorts.

https://adni.loni.usc.edu/data-samples/

https://wrap.wisc.edu/data-requests-2/

## Acknowledgements

We would like to thank all study participants, their supporters, and the study teams that contributed to this work.

Data collection and sharing for this project was funded by the Alzheimer’s Disease Neuroimaging Initiative (ADNI) (National Institutes of Health Grant U01 AG024904) and DOD ADNI (Department of Defense award number W81XWH-12-2-0012). ADNI is funded by the National Institute on Aging, the National Institute of Biomedical Imaging and Bioengineering, and through generous contributions from the following: AbbVie, Alzheimer’s Association; Alzheimer’s Drug Discovery Foundation; Araclon Biotech; BioClinica, Inc.; Biogen; Bristol-Myers Squibb Company; CereSpir, Inc.; Cogstate; Eisai Inc.; Elan Pharmaceuticals, Inc.; Eli Lilly and Company; EuroImmun; F. Hoffmann-La Roche Ltd and its affiliated company Genentech, Inc.; Fujirebio; GE Healthcare; IXICO Ltd.; Janssen Alzheimer Immunotherapy Research & Development, LLC.; Johnson & Johnson Pharmaceutical Research & Development LLC.; Lumosity; Lundbeck; Merck & Co., Inc.; Meso Scale Diagnostics, LLC.; NeuroRx Research; Neurotrack Technologies; Novartis Pharmaceuticals Corporation; Pfizer Inc.; Piramal Imaging; Servier; Takeda Pharmaceutical Company; and Transition Therapeutics. The Canadian Institutes of Health Research is providing funds to support ADNI clinical sites in Canada. Private sector contributions are facilitated by the Foundation for the National Institutes of Health (www.fnih.org). The grantee organization is the Northern California Institute for Research and Education, and the study is coordinated by the Alzheimer’s Therapeutic Research Institute at the University of Southern California. ADNI data are disseminated by the Laboratory for Neuro Imaging at the University of Southern California.

## Supplementary Material

See separate document.

## Abbreviations

AD: Alzheimer’s disease
*APOE*: apolipoprotein e
ETOA: estimated tau onset age
PET: positron emission tomography
SILA: sampled iterative local approximation
SUVR: standard uptake value duration

