## Supplemental Figure 1 for "Estimating tau onset age from tau PET imaging in two longitudinal cohorts using sampled iterative local approximation"

**Supplementary Materials**

**Figures**

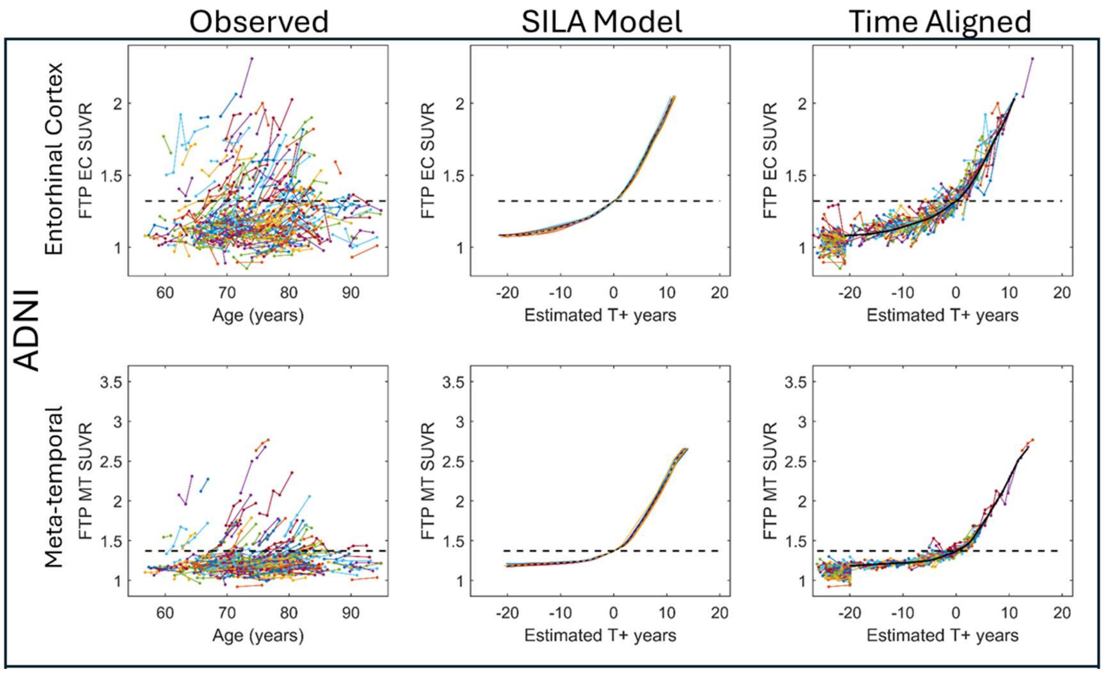


**Supplemental Figure S1:** ADNI observed and SILA-modeled tau PET SUVR trajectories excluding participants with Dementia (i.e., CDR ≥1)

**Supplemental Figure S2**

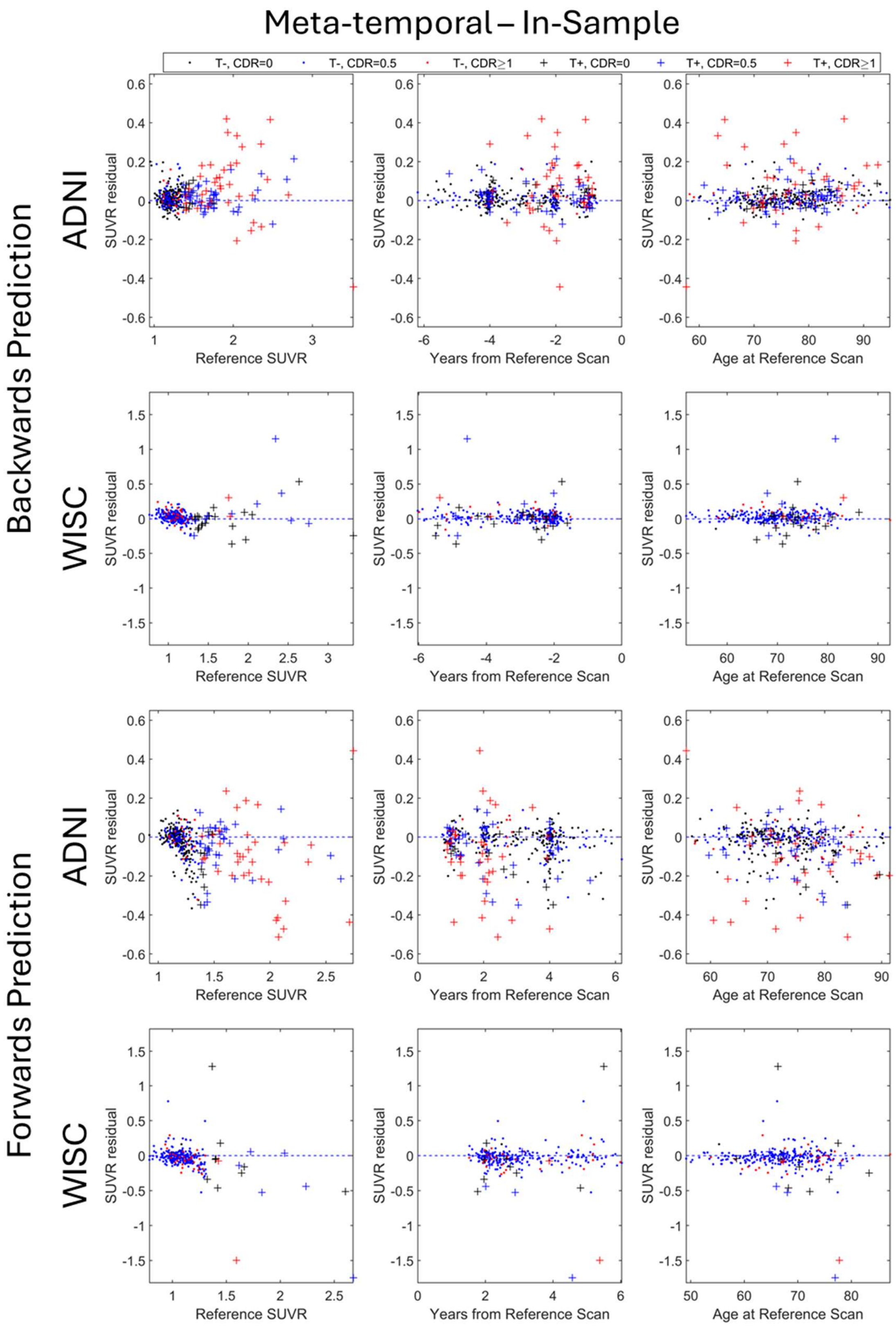

**Supplemental Figure S2:** Backward (top two rows) and forward (bottom two rows) SILA prediction residuals (observed – estimated) in the meta-temporal ROI from the in-sample training scheme as a function of age, reference SUVR, and time from reference scan. Color indicates CDR global score at the last available tau PET scan.

**Supplemental Figure S3**

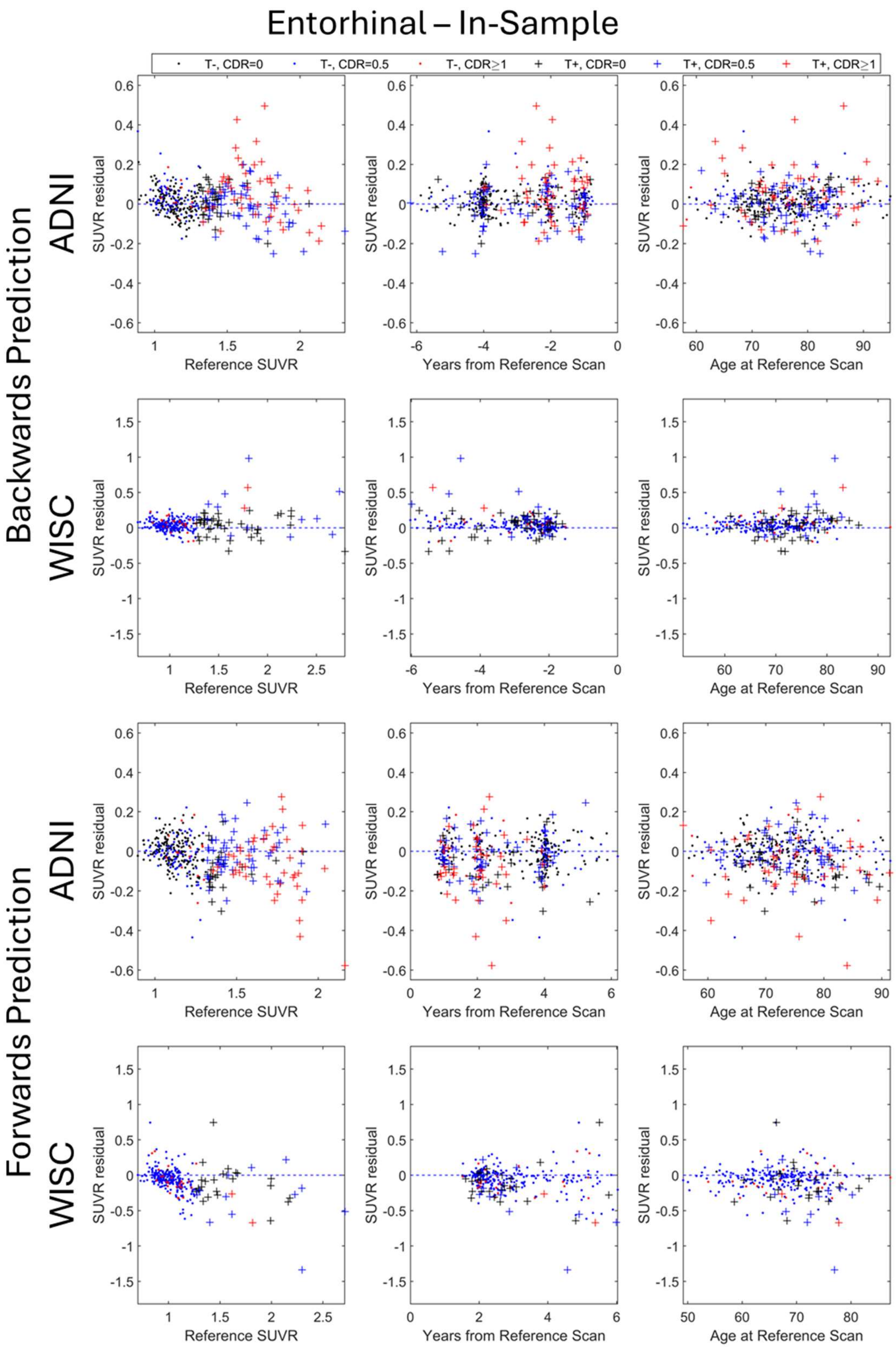

**Supplemental Figure S3** Backward (top two rows) and forward (bottom two rows) SILA prediction residuals (observed – estimated) in the entorhinal cortex ROI from the in-sample training scheme as a function of age, reference SUVR, and time from reference scan. Color indicates CDR global score at the last available tau PET scan.

**Tables**

**Supplemental Table S1. Ten-fold cross-validation summary statistics excluding participants with CDR ≥1 (ADNI only)**

| | | ADNI | |
|---|---|---|---|
| **Prediction Schema** | **Metric** | **Entorhinal** | **Meta-temporal** |
| **Backward** | **SSQ SUVR** | 18.27 | 12.12 |
| | **RMSE SUVR** | 0.055 | 0.037 |
| | RMSE(T+) | 0.082 | 0.063 |
| | RMSE(T-) | 0.049 | 0.033 |
| | **Bias SUVR** | 0.019 | 0.010 |
| | **T+/- Balanced Accuracy** | 82.6% | 86.1% |
| | **T+/- Accuracy** | 88.5% | 95.5% |
| | **Number of T- to T+ converters** | 24 | 17 |
| | **Conversion accuracy T- to T+, % ($n_{corr}$)** | 79.2% | 70.6% |
| | **Conversion midpoint error mean years [95% CI]** | -0.21 [-0.67, 0.25] | -0.35 [-0.86, 0.15] |
| **Forward** | **SSQ** | 24.63 | 17.19 |
| | **RMSE** | 0.075 | 0.052 |
| | RMSE(T+) | 0.095 | 0.074 |
| | RMSE(T-) | 0.068 | 0.047 |
| | **Bias SUVR** | -0.027 | -0.017 |
| | **T+/- Balanced Accuracy** | 86.3% | 93.4% |
| | **T+/- Accuracy** | 85.2% | 92.4% |

Model summary statistics for backward and forward SUVR, T+/- prediction and validation of estimated T+ onset age (ETOA) in a subset of observed T- to T+ converters (gray) using from 10-fold cross-validation results with dementia cases (CDR ≥1) excluded. Conversion accuracy was defined as the number of times ETOA was between the observed last T- and first T+ scan for converters. Conversion midpoint error was defined as the difference between ETOA and the midpoint between the last T- and T+ scan for observed converters.

**Supplemental Table S2. Ten-fold cross-validation summary statistics for partial-volume corrected data (ADNI only)**

| | | ADNI | |
|---|---|---|---|
| **Prediction Schema** | **Metric** | **Entorhinal** | **Meta-temporal** |
| **Backward** | **SSQ SUVR** | 45.94 | 30.63 |
| | **RMSE SUVR** | 0.120 | 0.080 |
| | RMSE(T+) | 0.197 | 0.160 |
| | RMSE(T-) | 0.099 | 0.056 |
| | **Bias SUVR** | 0.058 | 0.037 |
| | **T+/- Balanced Accuracy** | 81.7% | 92.4% |
| | **T+/- Accuracy** | 88.3% | 96.4% |
| | **Number of T- to T+ converters** | 33 | 18 |
| | **Conversion accuracy T- to T+, % ($n_{correct}$)** | 60.6% | 83.3% |
| | **Conversion midpoint error mean years [95% CI]** | 0.21 [-0.22, 0.64] | 0.29 [-0.04, 0.62] |
| **Forward** | **SSQ** | 56.04 | 40.01 |
| | **RMSE** | 0.146 | 0.104 |
| | RMSE(T+) | 0.223 | 0.205 |
| | RMSE(T-) | 0.120 | 0.067 |
| | **Bias SUVR** | -0.072 | -0.052 |
| | **T+/- Balanced Accuracy** | 85.6% | 92.7% |
| | **T+/- Accuracy** | 81.5% | 92.4% |

**Supplemental Table S3: Residual Associations with sex**
Residuals ~ Sex (Kruskal-Wallis Tests)
Cell: Absolute Difference (p-value)

| **Entire Dataset** | **ADNI EC** | **ADNI MT** | **WISC EC** | **WISC MT** |
|---|---|---|---|---|
| KF-forward | 0.008 (0.02) | 0.005 (0.19) | 0.002 (0.61) | 0.002 (0.19) |
| KF-backward | 0.004 (0.40) | 0.002 (0.77) | 0.003 (0.87) | 0.002 (0.74) |
| WH-forward | 0.010 (0.01) | 0.005 (0.23) | 0.002 (0.56) | 0.001 (0.24) |
| WH-backward | 0.002 (0.54) | 0.002 (0.90) | 0.0002 (0.90) | 0.003 (0.44) |
| **PVC** | | | | |
| KF-forward | 0.003 (0.65) | 0.002 (0.91) | - | - |
| KF-backward | 0.025 (0.01) | 0.010 (0.20) | - | - |
| WH-forward | 0.002 (0.72) | 0.004 (0.49) | - | - |
| WH-backward | 0.024 (0.03) | 0.010 (0.17) | - | - |
| **Dementia-free** | | | | |
| KF-forward | 0.008 (0.01) | 0.005 (0.23) | - | - |
| KF-backward | 0.004 (0.47) | 0.002 (0.55) | - | - |
| WH-forward | 0.010 (0.02) | 0.005 (0.17) | - | - |
| WH-backward | 0.002 (0.29) | 0.002 (0.41) | - | - |

**Supplemental Table S4: Residual Associations with *APOE-e4***
Residuals ~ e4+ (Kruskal-Wallis Tests)
Cell: Absolute Difference (p-value)

| **Entire Dataset** | **ADNI EC** | **ADNI MT** | **WISC EC** | **WISC MT** |
|---|---|---|---|---|
| KF-forward | 0.001 (0.22) | 0.008 (0.07) | 0.007 (0.34) | 0.008 (0.49) |
| KF-backward | 0.001 (0.78) | 0.003 (0.38) | 0.001 (0.86) | 0.004 (0.24) |
| WH-forward | 0.004 (0.04) | 0.007 (0.01) | 0.005 (0.76) | 0.004 (0.79) |
| WH-backward | 0.003 (0.18) | 0.002 (0.33) | 0.003 (0.32) | 0.004 (0.97) |
| **PVC** | | | | |
| KF-forward | 0.009 (0.68) | 0.004 (0.26) | - | - |
| KF-backward | 0.009 (0.13) | 0.007 (0.10) | - | - |
| WH-forward | 0.009 (0.29) | 0.004 (0.12) | - | - |
| WH-backward | 0.009 (0.35) | 0.007 (0.05) | - | - |
| **Dementia-free** | | | | |
| KF-forward | 0.006 (0.06) | 0.012 (0.002) | - | - |
| KF-backward | 0.004 (0.62) | 0.003 (0.21) | - | - |
| WH-forward | 0.006 (0.11) | 0.012 (0.001) | - | - |
| WH-backward | 0.004 (0.75) | 0.003 (0.13) | - | - |

**Supplemental Table S5: Residual Associations with Dementia**
Residuals ~ Dementia (Kruskal-Wallis Tests)
Cell: Absolute Difference (p-value)

| Entire Dataset | ADNI EC | ADNI MT |
|---|---|---|
| KF-forward | 0.032 (0.05) | 0.029 (0.002) |
| KF-backward | 0.024 (0.14) | 0.013 (0.02) |
| WH-forward | 0.026 (0.02) | 0.029 (0.02) |
| WH-backward | 0.022 (0.08) | 0.015 (0.05) |
| **PVC** | | |
| KF-forward | 0.044 (0.18) | 0.051 (0.19) |
| KF-backward | 0.032 (0.62) | 0.021 (0.66) |
| WH-forward | 0.049 (0.07) | 0.046 (0.81) |
| WH-backward | 0.037 (0.76) | 0.017 (0.96) |

**Supplemental Table S6: Residual Associations with Age**
Residuals ~ Age (linear mixed model)
Model: Residual ~ B_0 + B_1(Age) + b_oi(subject id) + e_ij
Cell: B_1 (95% bootstrapped CI)
p < 0.05 (*)

| Entire Dataset | ADNI EC | ADNI MT | WISC EC | WISC MT |
|---|---|---|---|---|
| KF-forward | -0.0008 (-0.002, -0.0001) * | -0.0016 (-0.0023, -0.0008) * | -0.0043 (-0.0062, -0.0025) * | -0.0030 (-0.0049, -0.0010) * |
| KF-backward | 0.0004 (-0.0006, 0.0007 | 0.0004 (-0.0001,0.0010) | 0.0002 (-0.0011,0.0015) | 0.0002 (-0.0010,0.0015) |
| WH-forward | -0.0080 (-0.0015, -0.0001) * | -0.0015 (-0.0023, -0.0008) * | -0.0039 (-0.0057, -0.0021) * | -0.0027 (-0.0045, -0.0009) * |
| WH-backward | 0.0002 (-0.0005, 0.0009) | 0.0004 (-0.0001,0.0010) | -0.000004 ( -0.0012, 0.0012) | -0.0001 (-0.0013, 0.0010) |
| **PVC** | | | | |
| KF-forward | -0.0023 (-0.0036, -0.0009) * | -0.0020 (-0.0031, -0.0008) * | - | - |
| KF-backward | 0.0002 (-0.0011, 0.0015) | 0.0006 (-0.0005, 0.0016) | - | - |
| WH-forward | -0.0023 (-0.0037, -0.0009) * | -0.0019 ( -0.0030, -0.0008) * | - | - |
| WH-backward | 0.0002 (-0.0011, 0.0015) | -0.0005 (-0.0005, 0.0015) | - | - |
| **Dementia-free** | | | | |
| KF-forward | -0.0010 (-0.0018, -0.0003) * | -0.0012 (-0.0018, -0.0006) * | - | - |
| KF-backward | -0.0003 (-0.0007,0.0006) | 0.0003 (-0.0001, 0.0007) | - | - |
| WH-forward | -0.0011 (-0.0018, -0.0004) * | -0.0012 (-0.0018, -0.0008) * | - | - |
| WH-backward | -0.00003 (-0.0006, 0.0006) | 0.0003 (-0.0001, 0.0007) | - | - |

**Supplemental Table S7: Residual Associations with reference tau PET SUVR**
Residuals ~ Reference SUVR (linear mixed model)
Model: Residual ~ B_0 + B_1(Ref. SUVR) + b_oi(subject id) + e_ij
Cell: B_1 (95% bootstrapped CI)
p < 0.05 (*)

| **Entire Dataset** | **ADNI EC** | **ADNI MT** | **WISC EC** | **WISC MT** |
|---|---|---|---|---|
| KF-forward | -0.076 (-0.10, -0.05)* | -0.07 (-0.09, -0.05)* | -0.17 (-0.21, -0.13)* | -0.27 (-0.03, 0.03)* |
| KF-backward | -0.004 (-0.02, 0.02) | -0.005 (-0.02, 0.01) | 0.01 (-0.02, 0.03) | 0.002 (-0.02, 0.03) |
| WH-forward | -0.05 (-0.07, -0.03)* | -0.08 (-0.09, -0.06)* | -0.16 (-0.20, -0.12)* | -0.25 (-0.31, -0.20)* |
| WH-backward | -0.02 (-0.04, 0.005) | 0.002 (-0.01, 0.02) | 0.02 (-0.004, 0.042) | 0.02 (-0.01, 0.04) |
| **PVC** | | | | |
| KF-forward | -0.076 (-0.10, -0.05)* | -0.10 (-0.13, -0.07)* | - | - |
| KF-backward | -0.004 (-0.02, 0.02) | -0.03 (-0.05, -0.002)* | - | - |
| WH-forward | -0.05 (-0.07, -0.03)* | -0.10 (-0.13, -0.07)* | - | - |
| WH-backward | -0.02 (-0.04, 0.005) | -0.02 (-0.04, 0.002) | - | - |
| **Dementia-free** | | | | |
| KF-forward | -0.08 (-0.11, -0.05)* | -0.05 (-0.07, -0.02)* | - | - |
| KF-backward | 0.01 (-0.01, 0.03) | -0.01 (-0.02, 0.01) | - | - |
| WH-forward | -0.08 (-0.10, -0..05)* | -0.04 (-0.06, -0.01)* | - | - |
| WH-backward | -0.01 (-0.01, 0.03) | -0.01 (-0.02, 0.001) | - | - |

**Supplemental Table S8: Residual Associations with Age**

Residuals ~ ETOA (linear mixed model)

Model: Residual ~ B_0 + B_1(ETOA) + b_oi(subject id) + e_ij

Cell: B_1 (95% bootstrapped CI)

$p < 0.05$ (*)

| Entire Dataset | ADNI EC | ADNI MT | WISC EC | WISC MT |
|---|---|---|---|---|
| KF-forward | 0.001 (0.001, 0.002)* | 0.001 (0.001, 0.002)* | 0.004 (-0.002, 0.005)* | 0.002 (0.001, 0.003)* |
| KF-backward | 0.0002 (-0.0002, 0.001) | 0.0002 (-0.0002, 0.001) | 0.001 (-0.003, 0.002) | 0.001 (-0.0002, 0.002) |
| WH-forward | 0.001 (0.0004, 0.001)* | 0.001 (0.001, 0.002) * | 0.004 (0.002, 0.005)* | 0.002 (0.001, 0.004)* |
| WH-backward | 0.001 (0.0001, 0.001) | 0.0001 (-0.0003, 0.0004) | 0.0004 (-0.001, 0.001) | 0.001 (-0.0003, 0.001) |
| **PVC** | | | | |
| KF-forward | 0.001 (0.001, 0.002)* | 0.002 (0.001, 0.003)* | - | - |
| KF-backward | 0.0002 (-0.0002, 0.001) | 0.001 (0.0003, 0.002)* | - | - |
| WH-forward | 0.001 (0.0004, 0.001)* | 0.002 (0.001, 0.003)* | - | - |
| WH-backward | 0.001 (0.0001, 0.001) | 0.001 (0.0003, 0.002)* | - | - |
| **Dementia-free** | | | | |
| KF-forward | 0.001 (0.0002, 0.001)* | 0.001 (0.001, 0.002)* | - | - |
| KF-backward | 0.0003 (0.00, 0.001)* | 0.0001 (-0.0003, 0.001) | - | - |
| WH-forward | 0.001 (0.0002, 0.001) * | 0.001 (0.001, 0.002)* | - | - |
| WH-backward | 0.0003 (0.0001, 0.001)* | 0.0001 (-0.0002, 0.001) | - | - |

**Supplemental Table S9: Residual Pearson Correlations**

| Cohort – Region | Variable | KF-Forward | KF-Backward | WH-forward | WH-backward |
|---|---|---|---|---|---|
| ADNI – ERC | Centered Age | -0.06 | 0.02 | -0.07 | 0.04 |
| | Female | 0.05 | 0.03 | 0.07 | 0.01 |
| | E4 Positive | 0.01 | 0.00 | 0.03 | -0.02 |
| | Dementia | -0.14 | 0.12 | -0.12 | 0.11 |
| | Reference SUVR | **-0.24** | -0.02 | **-0.17** | -0.11 |
| | ETOA | **0.19** | 0.03 | 0.13 | 0.10 |
| ADNI – Meta-temporal | Centered Age | -0.13 | 0.09 | -0.13 | 0.09 |
| | Female | 0.03 | 0.02 | 0.03 | 0.02 |
| | E4 Positive | 0.05 | -0.03 | 0.05 | -0.02 |
| | Dementia | -0.13 | 0.08 | -0.13 | 0.09 |
| | Reference SUVR | **-0.25** | -0.03 | **-0.28** | 0.01 |
| | ETOA | **0.19** | 0.04 | **0.20** | 0.01 |
| WISC - ERC | Centered Age | -0.18 | 0.05 | -0.17 | 0.03 |
| | Female | -0.01 | -0.01 | -0.01 | 0.00 |
| | E4 Positive | -0.03 | 0.01 | -0.02 | 0.02 |
| | Dementia | - | - | - | - |
| | Reference SUVR | **-0.33** | 0.02 | **-0.33** | 0.07 |
| | ETOA | **0.24** | 0.07 | **0.23** | 0.04 |
| WISC – Meta-temporal | Centered Age | -0.13 | 0.03 | -0.13 | 0.01 |
| | Female | -0.01 | -0.01 | -0.01 | -0.01 |
| | E4 Positive | -0.03 | 0.03 | -0.02 | 0.03 |
| | Dementia | - | - | - | - |
| | Reference SUVR | **-0.40** | -0.003 | **-0.40** | 0.05 |
| | ETOA | 0.13 | 0.08 | **0.18** | 0.06 |

***Boldface** indicates $p<0.05$

**Supplemental Table S9: Residual Pearson Correlations for ADNI partial-volume corrected SUVR**

| Cohort – Region | Variable | KF-Forward | KF-Backward | WH-forward | WH-backward |
|---|---|---|---|---|---|
| ADNI – ERC | Centered Age | -0.09 | 0.04 | -0.09 | 0.04 |
| | Female | -0.01 | 0.09 | -0.01 | 0.09 |
| | E4 Positive | -0.03 | 0.04 | -0.03 | 0.04 |
| | Dementia | -0.11 | 0.08 | -0.12 | 0.10 |
| | Reference SUVR | **-0.15** | 0.03 | **-0.17** | 0.05 |
| | ETOA | **0.16** | 0.07 | **0.18** | 0.04 |
| ADNI – Meta-temporal | Centered Age | -0.10 | 0.08 | -0.11 | 0.08 |
| | Female | -0.01 | 0.05 | -0.02 | 0.05 |
| | E4 Positive | 0.02 | -0.03 | 0.02 | -0.03 |
| | Dementia | **-0.15** | 0.07 | -0.14 | 0.06 |
| | Reference SUVR | **-0.22** | -0.09 | **-0.24** | -0.07 |
| | ETOA | **0.15** | 0.11 | **0.16** | 0.11 |

**Supplemental Table S10: Residual Pearson Correlations for ADNI SUVR without dementia cases**

| Cohort – Region | Variable | KF-Forward | KF-Backward | WH-forward | WH-backward |
|---|---|---|---|---|---|
| ADNI – ERC | Centered Age | -0.09 | 0.02 | -0.10 | 0.02 |
| | Female | 0.05 | 0.01 | 0.05 | 0.01 |
| | E4 Positive | 0.04 | 0.04 | 0.05 | 0.03 |
| | Reference SUVR | **-0.23** | 0.03 | **-0.23** | 0.02 |
| | ETOA | **0.22** | 0.02 | **0.21** | 0.03 |
| ADNI – Meta-temporal | Centered Age | -0.14 | 0.08 | -0.15 | 0.09 |
| | Female | 0.07 | -0.05 | 0.07 | -0.06 |
| | E4 Positive | 0.11 | -0.04 | 0.12 | -0.04 |
| | Reference SUVR | **-0.17** | -0.02 | -0.14 | -0.06 |
| | ETOA | 0.14 | 0.09 | 0.13 | 0.10 |
